# COVID-19’s U.S. Temperature Response Profile

**DOI:** 10.1101/2020.11.03.20225581

**Authors:** Richard T. Carson, Samuel L. Carson, Thayne Keegan Dye, Samuel A. Mayfield, Daniel C. Moyer, Chu A. (Alex) Yu

## Abstract

We estimate the U.S. temperature response curve for COVID-19 and show transmission is highly sensitive to temperature variation despite summer outbreaks widely assumed to show otherwise. By largely replacing the daily death counts states initially reported with counts based on death certificate date, we build a week-ahead statistical forecasting model that explains most of the daily variation (R^2^ = 0.97) and isolates COVID-19’s temperature response profile (p < 0.001). These counts normalized at 31°C (U.S. mid-summer average) scale up to nearly 160% at 5°C. Positive cases are more temperature sensitive, scaling up to almost 400% between 31°C and 5°C. Dynamic feedback amplifies these effects, suggesting that cooling temperatures are likely to the substantially increase COVID-19 transmission.

**Article Summary Line:** COVID-19’s temperature response profile is reliably estimated using re-assembled state-reported data and suggests the onset of cold weather will amplify its spread.

## Introduction

The question of whether COVID-19 exhibits a pronounced temperature response profile has garnered attention since the early days of the pandemic *(1,2,3)*. Kissler et al.’s examination of medium and long-term management of the pandemic assigns a prominent role to understanding how the temperature response profile (hereafter “TRP”) of COVID-19 might influence the pandemic’s progression in the United States *(3,4)*. This follows conventional wisdom regarding the strong seasonal pattern of influenza *(5,6)*, which helped mask COVID-19’s early U.S. ascent *(7)*. The recent rise in positive COVID-19 cases and related deaths across the United States has called into question whether COVID-19 transmission is adversely sensitive to summer weather. Nevertheless, some modeling groups such as the University of Washington’s Institute for Health Metrics (IHME) have assumed COVID-19 activity will increase as temperatures decline and are now using information from the U.S. influenza monitoring network *(8)*, which was previously shown predictive of the pandemic’s path last spring *(7)*, to help incorporate that effect.

Daily state reports often bear little resemblance to the number of COVID-19-related deaths actually occurring on that date. The resulting temporal data misalignment creates a substantial impediment to recovering any relationships with critical dependence on event timing. By reconstructing the death counts states report daily – largely by substituting in retroactive corrections based on death certificate dates – we are able to reliably estimate the TRP for COVID-19 deaths.

We assemble over 2,500 state-level daily observations from April 16–July 15, 2020, after COVID-19 became well-established across the United States. We follow the environmental economics literature on estimating pollution and temperature related impacts on a range of health and other outcomes (9). SARS-CoV-2 being a novel virus makes full implementation of the standard approach infeasible because multiple years of both spatially and temporally delineated data are not available. The specific issue we cannot currently resolve is whether the TRP over a given temperature range is the same in both directions, i.e. the cooler-to-warmer direction reflected in our data set and the warmer-to-cooler direction now occurring. It is also important to recognize that we estimate the joint, not separate, effect of any biological response by the virus and any behavioral response by the public.

### Prior Efforts

Early attempts to pin down COVID-19’s temperature response profile (TRP) proved it to be elusive. The now well-accepted approach for estimating influenza’s TRP is epitomized by Barreca and Shimshack *(6)*, which draws heavily on the modelling of climate impacts on human populations *(9,10,11)*. Under this approach a panel data set of political entities, such as countries or their political subregions (states, counties, etc.), is assembled and the outcome of interest observed across a long time horizon (e.g., a 20-year period). The ability to employ fixed-effect indicator variables to correct for time-invariant differences between political jurisdictions, coupled with the use of short-run weather variability, provides statistical identification of a variety of impact response functions.

Prior research has three important limitations. First, cross-sectional data cannot statistically identify the desired function without making the implausible assumption that all possible confounding variables are adequately controlled for. Routinely updated time-series models slowly incorporate environmental conditions into their forecasts without ever isolating it. Short panel datasets, where the stimulus of interest has limited range (temperature, humidity, UV light, air pollution in each location), often lack the statistical power to pin down such response functions. Consequently, estimated response functions are often fragile in the sense that statistical significance is lost when time trends or demographic variables are added to models *(12,13,14,15)*.

Second, early work (often using Chinese or cross-country data) focused on predicting speed at which the pandemic ramped up in different locations, using variants of derivative statistics like growth rates or R0*(16,17)*. These works point the 0-10°C temperature range as being most conducive to spreading COVID-19, with a possible humidity effect *(17,18,19)*. Current interest is now focused on situations where COVID-19 is spatially well-seeded and its effective R0 can move up or down with actions like state reopening plans.

Third, the quality and comparability of reported COVID-19 statistics is often suspect, particularly from the early phase of the pandemic. We move past this period to an observational window where reporting largely stabilized. A different problem now dominates temporal mismatches between when an event (e.g., a COVID-19-related death) was reported and the weather variables potentially influencing that event *(11)*. When reported event dates significantly differ from actual event dates, the resulting measurement error can overwhelm standard sources of biological variation such as individual differences in incubation periods.

### Correcting State-Level COVID-19 Statistics And Why It Matters

Our ability to isolate the TRP for COVID-19 related deaths stems largely from our reconstruction of state-level COVID-19 data. The most important of these is replacing the daily death counts initially reported by states with the revised daily counts based on actual death certificate date where possible. If death certificate data is not available, we use the retroactively corrected data series that several states have produced. When available, this type of data generally rectifies many initial reporting errors. We also correct other implausible data reports such as implicit negative daily death counts and zero counts on one day followed by a clear double-count the following day. The Appendix Data Preparation section provides details.

Figure 1 displays why repairing daily state-level COVID-19 death counts matters. Panel shows the originally reported (COVID Tracking Project [CTP]) CTPDailyDead_it_ daily death counts for (Florida) in blue with the “Revised” death counts by death certificate date overlaid in red. Revised curves follow the general shape predicted by epidemic models, whereas curves based on the originally reported counts have clear day-of-the-week patterns and large spikes, neither of which are predicted by biological models. Panel (B) shows the two implications. First, the confidence interval from fitting a simple quadratic trend model is dramatically larger for the CTPDailyDead_it_ (i.e., the Reported data) than the same model fit using the Revised. Second, the model based on the CTPDailyDead_it_ is slow to pick up the sharp rise in Florida deaths because the originally reported counts temporally lag the revised counts. Figure 1, Panels (C) and (D) display results for Georgia which show how use of CTPDailyDead_it_ can lead to both missing a downturn and an upturn, while supporting an incorrect story of slow progress. Appendix Figure A1 provides similar graphs for Arizona and Texas.

**Figure 1.**
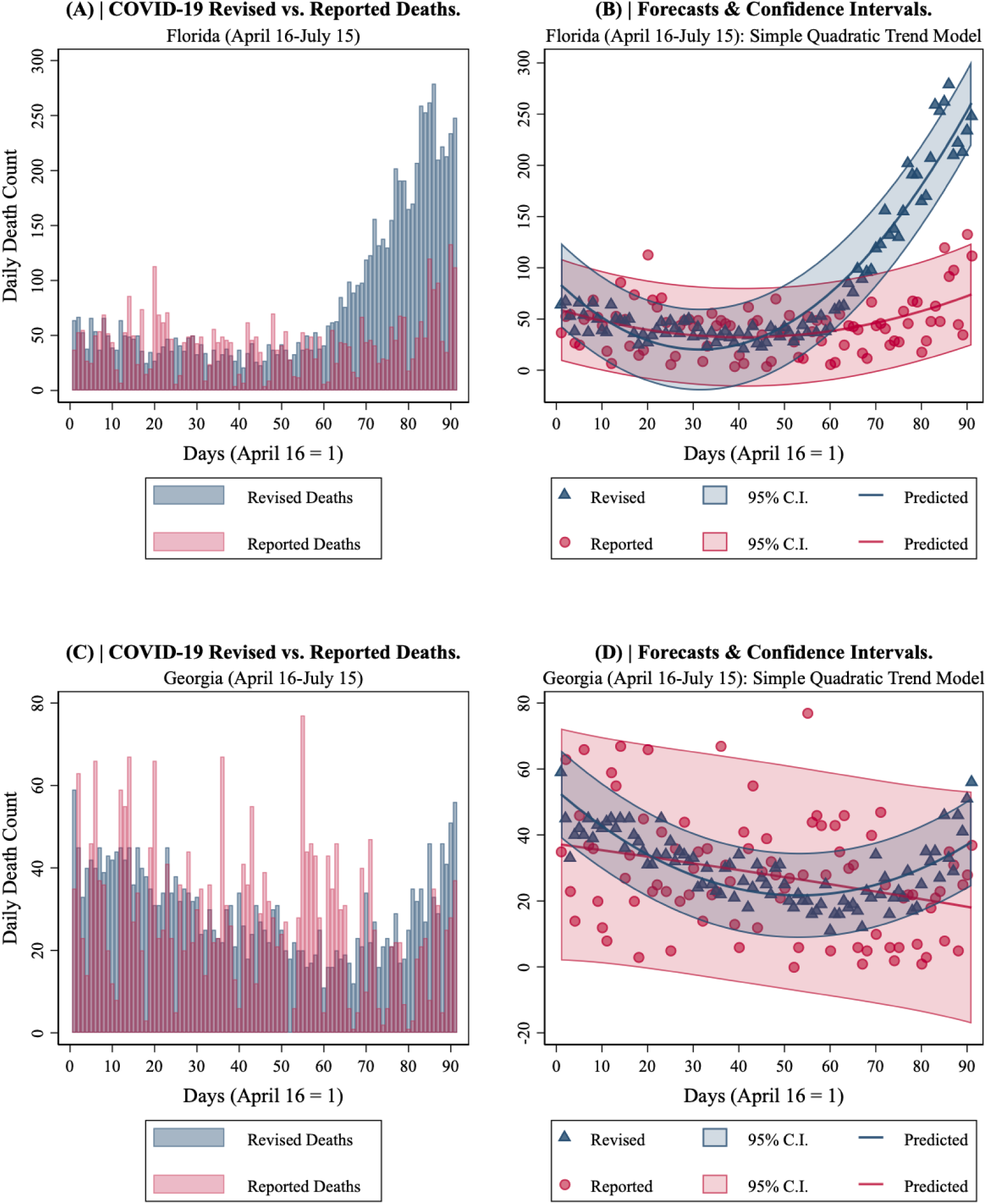
COVID-19 Deaths in Florida and Georgia. April 16-July 15. **(A)** Florida Revised vs. Reported. **(B)** Florida quadratic time trend forecasts and confidence intervals. **(C)** Georgia Revised vs. Reported. **(D)** Georgia quadratic time trend forecasts and confidence intervals.

Figure 2(A) shows the week ahead forecast from the University of Washington’s Institute of Health Metrics and Evaluation (UW-IHME) plotted against our model’s DailyDead_it_ at the state level for the entire United States over the three-month period we examine. This model explains a reasonable amount of the variance, reflected in the R^2^ of .66 obtained by regressing the Revised on their forecasts. Substituting in the week-ahead forecasts from one of the other heavily used forecasting models results in a similar overall impression.

**Figure 2.**
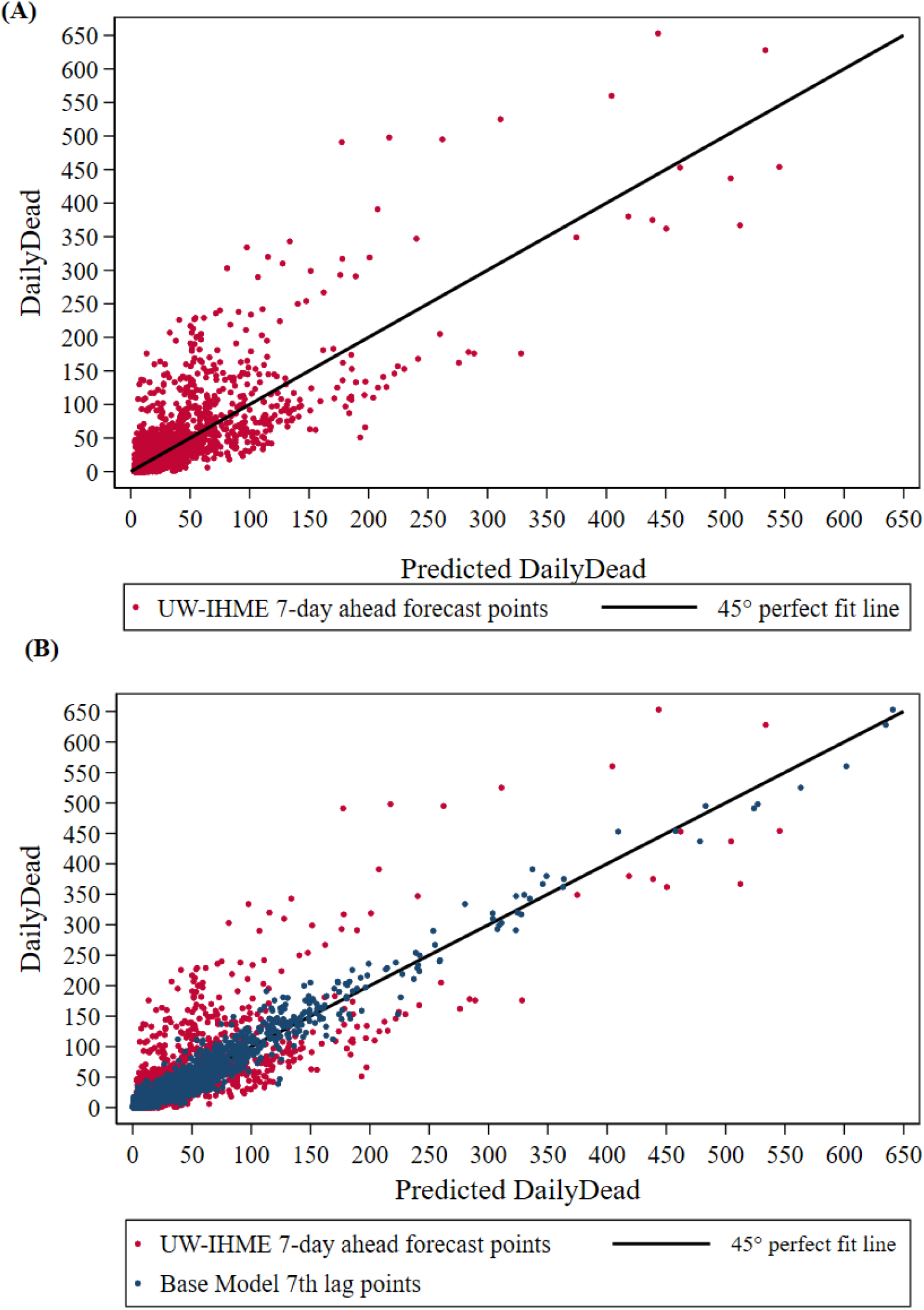
DailyDead_it_ vs. predicted values using information available 7 days earlier. U.S. States April 16-July 15. **(A)** UW-IHME forecasts (R^2^ = .66). **(B)** Eq. 1 base model predictions (R^2^ = .97).

Figure 2(B) plots the predicted DailyDead_it_ using Eq. 1, which only uses information available a week earlier. This regression’s R^2^ of .97 is reflected in the very tight scatter plot. Since our model is relatively simple, the main source of the improvement in the percent of the variance explained is building the model off the temporally-aligned death counts provided by states post-hoc.

### Modelling Approach

Our objective is to isolate COVID-19’s TRPs with respect to state-level daily death counts (DailyDead_it_) and new positive test counts (NewPositives_it_). Any TRP specification should allow for the possibility of non-responsiveness (i.e., zero temperature dependence). TRPs should not incorporate fixed factors like state demographics, nor other factors associated with calendar date or a clear temporal profile. Rather, daily exogenous variation in temperature on any specific day should be the source of statistical information for identifying the TRP of interest. Intuitively, the TRP is being statistically identified by having days where the lagged count of the COVID-statistic of interest is approximately the same magnitude but a range of different temperatures is observed. The number of such comparable days is substantially increased by the introduction of controls for conditions that remain fixed across states and a flexible time trend. Statistical modeling issues revolve around functional forms and the specification of the relevant lag structure. The slow-moving, systematic changes in temperature over time imply that, over short time horizons, temperature will not be the driving force behind DailyDead_it_ and NewPositives_it_. However, over longer time horizons, a virus’s TRP can be a major factor.

We index DailyDead_it_ and NewPositives_it_ by state (*i*) and day (*t*). For each we build a simple week-ahead forecasting model, controlling for state-level fixed effects and including a quadratic time trend. We then examine how lagged (t-k) maximum daily temperature (MaxTemp_it-k_) scales the model’s predicted DailyDead_it_ or NewPositives_it_. Attention is restricted to conditions where MaxTemp_it_ ≥ 5°C, since data below this level is sparse during our sample period and concentrated in a few sparsely populated states like Alaska and Montana. Empirical estimates of our TRPs are normalized to 100 at 31°C (∼88°F) – the U.S. population-weighted average for the last week of our sample – to aid interpretability.

The pandemic modelling community has largely concentrated on DailyDead_it_, believing it to be a more reliable indicator of the COVID-19 infection pool than NewPositives_it_ due to the large differences in testing regimes across states and time. We proceed in a similar manner, but also produce the TRP for NewPositives_it_ (conditioning on available testing information) since it is positive cases that are potentially directly influenced by temperature.

### Model Components

Our base DailyDead_it_ statistical model (Eq. 1) is comprised of two multiplicative components. The first produces expected current-period deaths as a function of past observed deaths at a fixed temperature. The second allows expected DailyDead_it_ to (potentially) vary with past values of MaxTemp_it-k_.

The terms inside the first component are the infection pool proxy, DailyDead_it-7_, state-level fixed effect indicators, StateIndicator_i_ and a quadratic time trend in Days_t_ (t=1, …, 137; initialized to March 1 to aid interpretability). The StateIndicator_i_ captures the influence of a wide range of variables which remain constant over the period examined, such as demographic composition, geographic links between locales that influence infections, and public health infrastructure. The time trend variables pick up the decline in the case fatality rate and the average effect over time of initial lockdowns, social distancing, and state re-openings. The first component is exponentiated to incorporate the restriction that expected DailyDead_it_ should not be negative if COVID-19 transmission is active anywhere in the set of connected units examined. Commensurately, we use LogDailyDead_it-k_ and LogDays_t_ as regressors, so this component can be interpreted as a log-log regression model with state-level fixed effects.

The second component is a logistic function scaling predicted DailyDead_it_ up or down with MaxTemp_it-k_. Deaths on specific days are the result of infections propagated over earlier days. We use LogMaxTemp_it-7_ and LogMaxTemp_it-14_ (one and two weeks in the past respectively) to roughly encompass the relevant period for temperature influencing current period deaths. Our panel data model specification uses an additive error term necessitates using nonlinear least squares to solve the model (*20,21*), but decouples conditional mean estimates from the estimated error component.

Formally, our base model for U.S. state *i* on day *t* is given by:

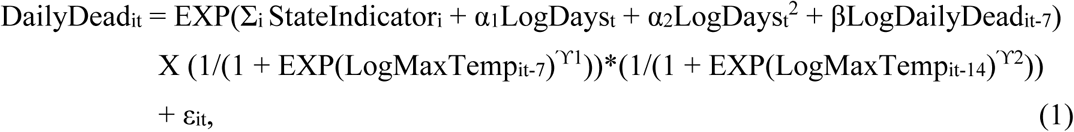

where estimated coefficients for the StateIndicator_i_, and the Greek letter parameters minimize the sum of the squared error term of the estimated ε_it_. The NewPositives_it_ model is similarly structured.

We investigate (Appendix Table A6) the sensitivity of our results to a range of alternative specifications (e.g., different infection pool indicators, different temperature scaling functions, adding absolute humidity, relative humidity, ultraviolet radiation, inclusion of shelter-in-place/reopening orders, lagged cumulative death counts, and use of the CTP death counts) and provide further discussion of modelling issues in the Appendix Modeling Approach section.

## Data

Our analysis uses three main types of data:

1. COVID-19 statistics for state-level death counts, positive cases, and tests, using The COVID Tracking Project (CTP; covidtracking.com) as our base information source.
2. Temperature data from the U.S. National Weather Service Integrated Surface Database.
3. State-level indicators and time variables.

We undertook extensive repair of the COVID-19 data reported daily by states, particularly those involving death counts. A dominant feature of these data are the substantial lags between when many of these events officially occurred and when they are reported. It is not uncommon to see states include deaths from several weeks prior in any given day’s count.

Over half of the U.S. states have made the number of COVID-related deaths publicly available by their death certificate dates in some manner (Appendix, Data Preparation). A simple OLS regression of death counts by death certificate date on originally reported death counts for many of these states yields an R^2^ of less than 0.5. Other states have made corrections to originally reported COVID-19 statistics. These updated counts tend to retroactively correct a myriad of reporting errors by states contained in the CTP daily data snapshot. These issues include failing to report any information on specific days, decisions on “probable” COVID-related deaths, and the resolution of duplicated death certificates. We use states’ self-corrected counts when available.

When neither of these two sources of information was available, we undertook a consistent set of data repair operations. These include: averaging across days with no reporting, prorating backwards in time large batches of deaths (and other statistics) known to have occurred over a much longer period (typically from nursing homes), and making the minimum set of changes necessary to correct logical violations such as negative counts of new deaths, positive cases, and tests. We make this dataset available for use by other researchers, along with a line-by-line account of the corrections made.

For each state, weather variables are taken from the airport with the highest volume of commercial traffic. We focus on maximum daily temperature. Mean temperature results are quite similar. State-level aggregation requires taking weather data from a single station. Measurement error induced by this compromise is likely to be less than one might think. Many states are small spatially or have a single, concentrated metropolitan area (e.g., Illinois). In spatially large states with large populations, most of the population often lives within reasonable proximity to the largest airport. Even in Texas, most people live along the corridor between Dallas (DFW is our representative airport) and Houston. As a result, over 60% of the American population lives within 300km of the representative airport for their state. Similarly, positive cases are also concentrated in major metropolitan areas near those airports. More generally, a single source of classical measurement error attenuates parameter estimates toward finding no effect.

### Daily Dead Model Results

Eq. 1 is estimated using non-linear least squares and has an R^2^ of 0.97. All parameter estimates (provided in Appendix Table A1) using robust standard errors clustered at the state-level are significant. Figure 2(B) displays the Revised DailyDead_it_ versus the model’s in-sample predicted values. States where the actual death certificate dates were available have considerably smaller prediction errors (Model 2 in Appendix Table A6).

The model’s quadratic time trend suggests DailyDead_it_ has fallen over time; at a declining rate from mid-April through the end of May, and then almost flat (slowly rising). LogDailyDead_it-7_ is the dominant predictor and its coefficient estimate of 0.8686 (t=35.32) has a standard elasticity interpretation.

Figure 3 shows the estimated TRP implied by the parameter estimates for the two MaxTemp_it_ lags in the logistic scaling function. The vertical axis represents expected DailyDead_it_ at each temperature value when both lags are set to the same temperature and the TRP is normalized to 100 at 31°C. (Appendix Figure A9 show the corresponding TRP using the original CTP death counts.) Appendix Figure A2 provides the contour plot which allows the two MaxTemp_it-k_ lags values to vary independently.

**Figure 3.**
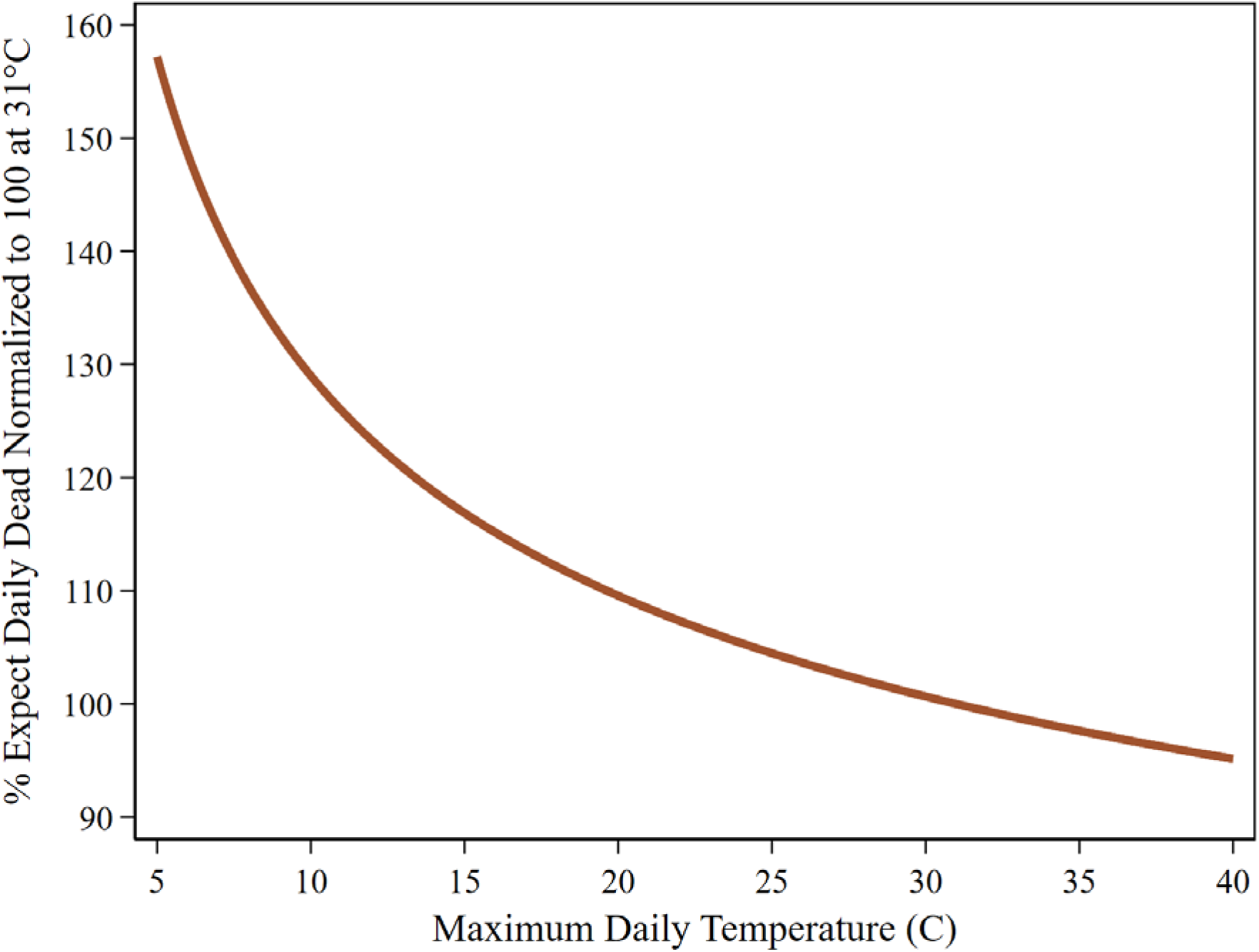
COVID-19 daily dead temperature response profile. U.S. states: April 16-July 15. Temperatures for 7th & 14th lags set equal.

Differentiating Eq. 1 with respect to DailyDead_it-7_ produces a measure of how the expected DailyDead_it_ increases from a one death increase in DailyDead_it-7_. Setting *t* and MaxTemp_it-k_ to chosen values yields [TRP*β*EXP(StateBase_it_)]/(DailyDead_it-7_)^(1-β)^, where the TRP directly scales the elasticity parameter, β, on LogDailyDead_it-7_, and StateBase_it_ is the temporally varying sum of the fixed StateIndicator_i_ and the quadratic time trend. As an example, for Georgia on July 15 (StateBase_it_ = 3.9184, MaxTemp_it-7_ = 29.4, MaxTemp_it-14_ = 27.8 and the corresponding non-normalized TRP = 0.0368), changing DailyDead_it-7_ from 27 to 28 is predicted to increase the expected DailyDead_it_ by 1.0457. Appendix Figure A3(B) displays a variant of this calculation for individual states as DailyDead_it-7_ increases from 0 to 1.

We simulate DailyDead_it_ predictions from both static and dynamic variants of our estimated base model (Eq 1) by taking DailyDead_it-7_ from our sample period’s last week and MaxTemp_it_ = 31°C as the initial values to propagate the simulation. We fix MaxTemp_it-7_ and MaxTemp_it-14_ at 31°C from day 1 to day 45 to mimic the rest of the summer period, then progressively decrease them by 0.2°C each day until 5°C is hit, which occurs on day 175, after which temperature is held constant.

The brown dashed line in Figure 4 provides a stylized static [left vertical axis] representation of information contained in our base model (Eq. 1) using Georgia as an example (since it starts out at just below our 31C° normalization point and in a cold year hits our 5C° endpoint). The two MaxTemp_it-k_ change in tandem, with all other variables fixed at their initial values. In this static response model expected DailyDead_it_ increases through only one channel: the direct impact of lowering temperature.

**Figure 4.**
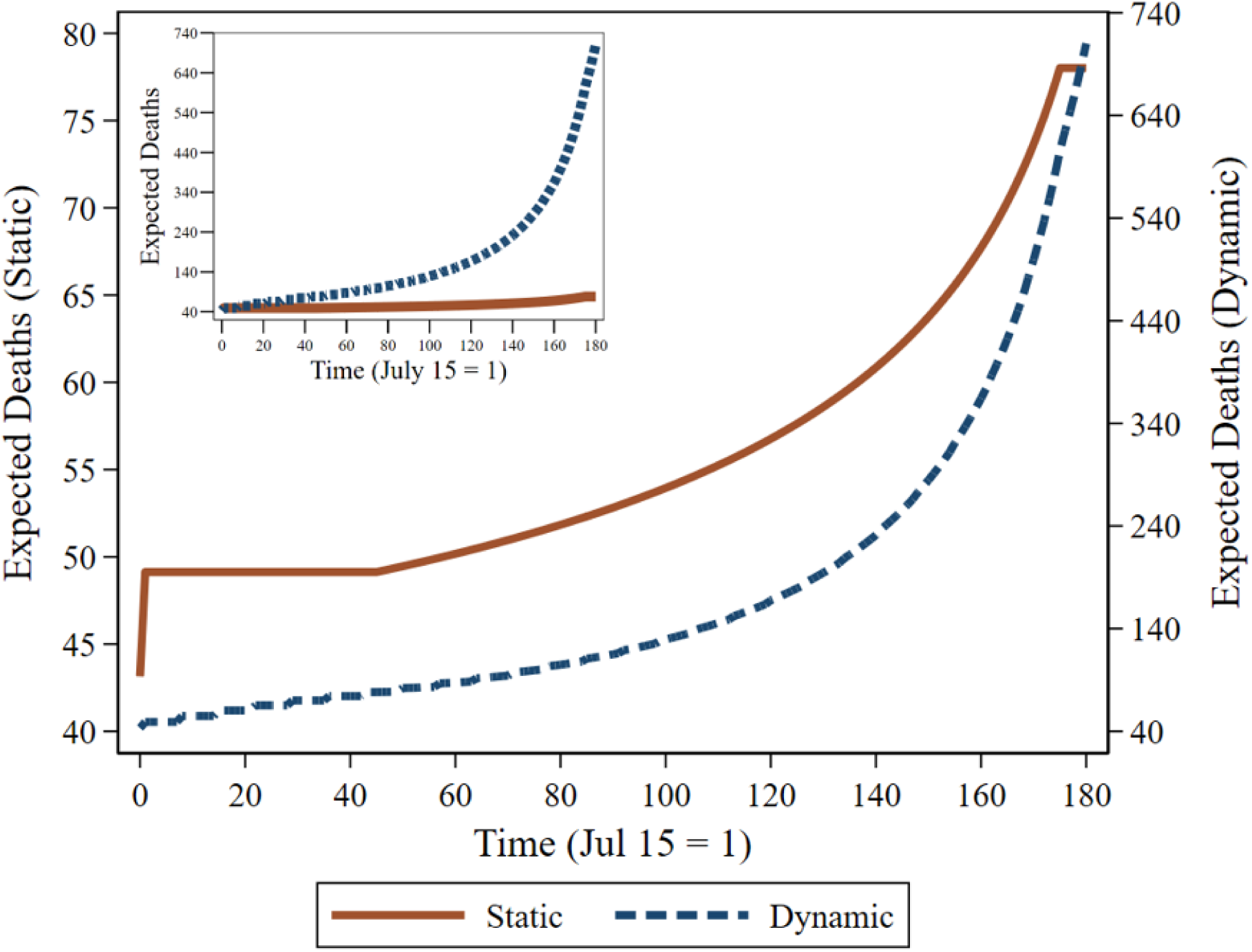
Stylized static and dynamic TRPs for Georgia derived from Eq. 1 parameter estimates. The brown solid curve [left vertical axis] is the stylized static TRP where temperature influences DailyDead_it_ through changes to MaxTemp_it-k_, with DailyDead_it-7_ held constant. The blue dashed curve [right vertical axis] is stylized dynamic TRP under similar assumptions, except dynamic feedback is allowed in the form of temperature influencing subsequent DailyDead_it-7_.

The blue dashed curve in Figure 4 shows the dynamic counterpart [right vertical axis] that allows MaxTemp_it-k_ to influence DailyDead_it_, which is then used as the lagged model input for subsequent projections. Temperature affects expected DailyDead_it_ through two channels: the direct effect shown in the static model and the indirect, compounding effect of when the initial death count of each cycle of the simulation is set to a lag of the previous output. The indirect effect of maximum temperature is the dominant mechanism, accounting for more than 90% of the increase in the expected death count as temperature falls from 31°C to 5°C. The inset in Figure 4 plots the two curves under the same scale and conveys the magnitude of the differences between the pure static and dynamic responses.

Neither model is likely to represent reality, but together may span it. Staying on the static TRP curve requires continual reductions in effective contact rates that exactly offset the increase in transmission potential, such that the current death count is always held to be equal to last week’s death count. Observing the full dynamic effect would require the absence of offsetting actions by both the government and the public (such as increased social distancing and face mask adherence as COVID-19 activity quickly increased).

StateIndicator_i_ values for some geographically isolated states with small populations like Hawaii are small enough to suggest their COVID-19 death counts would be unsustainable in warm enough weather. This is not true of most states, though, and is inconsistent with Baker et al.’s finding from examining earlier emergent viruses that warming weather is not enough by itself to stop their spread *(22)*.

The Appendix section on Alternative Specifications for Base Death Count Model describes additional analyses that (a) look at alternative temperature scaling functions, (b) substitute DailyDead_it-14_ or NewPositives_it-7_ for DailyDead_it-7_, and (c) add the dates of state actions such as shelter-in-place orders as control variables. This work shows that our finding of a strong TRP for DailyDead_it_ is quite robust. Some specifications suggest the TRP is flatter in the 10-20°C range but steeper in the 5-10°C range (Appendix Figure A8).

### New Positive Case Model Results

NewPositives_it_ are modeled similarly to Eq. 1, substituting LogNewPositives_it-7_ as the infection pool regressor. Testing information is required to interpret reported state-level positives cases. We use LogNewTests_it_, to control for current testing intensity, LogNewTest_it-7_ (which allows for a past positivity rate interpretation), total tests administered per thousand lagged by one week (PerCapitaTests_it-7_) to help control for prior testing intensity, and an indicator variable for systematically lower reporting on Monday. We chose LogMaxTemp_it-7_ for consistency with the death count model. For the other temperature lag the 2^nd^ lag fits best.

This model’s (Appendix Table A2) R^2^ is 0.95. All regressors are significant at p < 0.001, except for some of the test related variables: LogNewTest_it-7_ (p=0.005), PerCapitaTests_it-7_ (p=0.002) and Monday (p=0.015). Estimated parameters for the two lagged temperature variables are considerably larger than their DailyDead_it_ counterparts. The implied TRP for NewPositives_it_ is displayed in Figure 5. Appendix Figure A(4) shows contour plot allowing the two temperature lags to vary independently.

**Figure 5.**
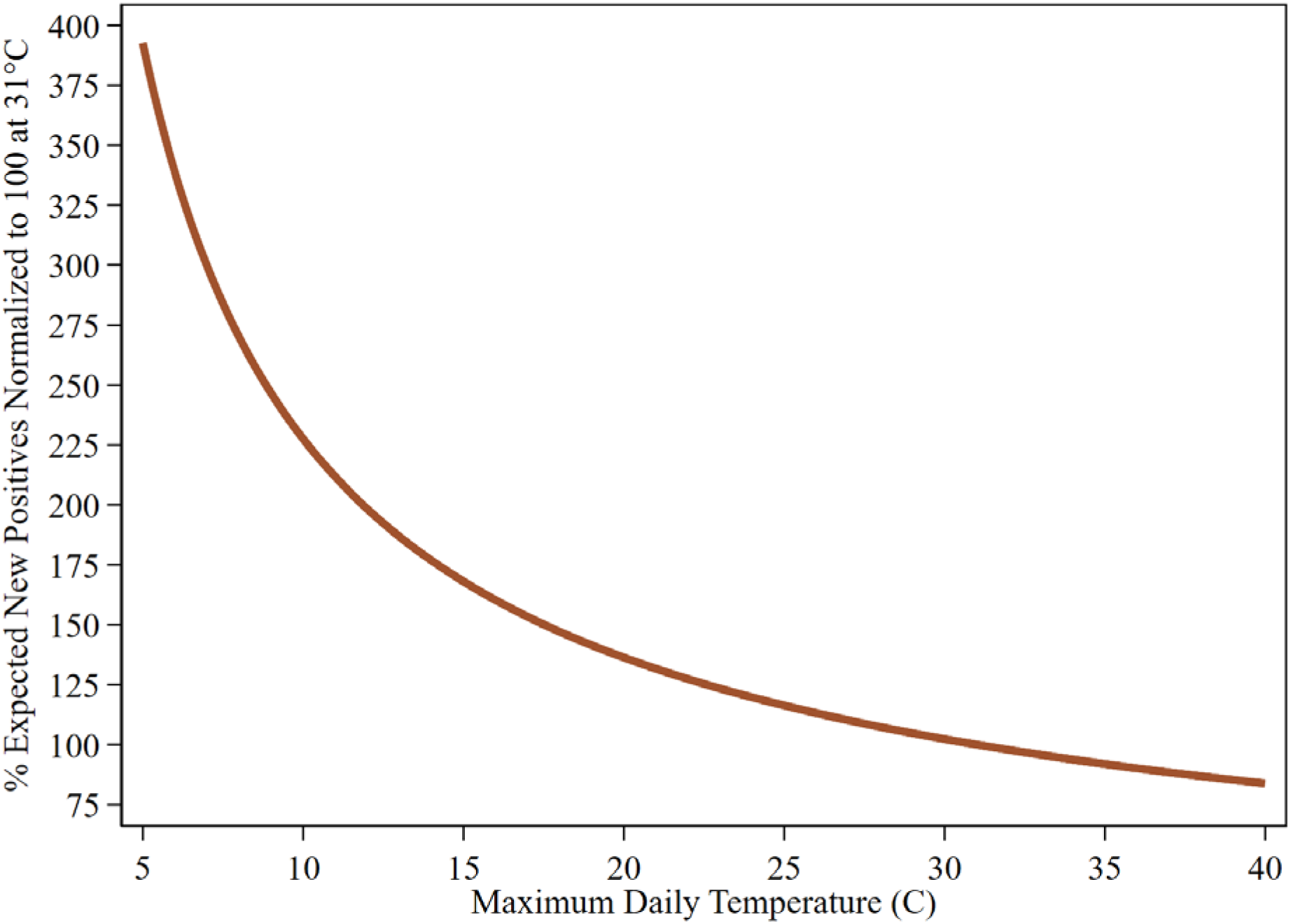
COVID-19: new positives temperature response profile. U.S. states: April 16-July 15. Temperatures for 2nd and 7th lags set equal.

## Discussion

DailyDead_it_ predictably varies with changes in maximum daily temperature (Figure 3). This relationship is considerably more pronounced for new positive cases (Figure 5). Cooler temperatures with the progression of fall and winter will dramatically ramp up the number of new positive cases and the deaths that follow unless current infection pools are dramatically reduced. Dynamic feedback between rising infection pool indicators and cooling temperatures (Figure 4) suggests delay in responding to increased virus activity signs can result in rapid escalation. This is already being seen in the current spatial pattern of outbreaks and the ever-increasing medium-run death count forecasts (e.g., UW-IHME *(8)*). Investment in providing the pandemic modeling community with timely counts based on death certificate dates would allow them to deliver substantially more accurate and timely warnings of impending upturns.

Warming temperatures during the spring and summer actively aided efforts to reduce COVID-19’s spread and may have even contributed to a false sense of the efficacy of those efforts. Cooling temperatures will present a quite different challenge. Figure 6 shows the average date over the past 30-years when each U.S. county enters the particularly dangerous 10°C to 5°C. Effects of temperature amplification show up with a lag.

**Figure 6.**
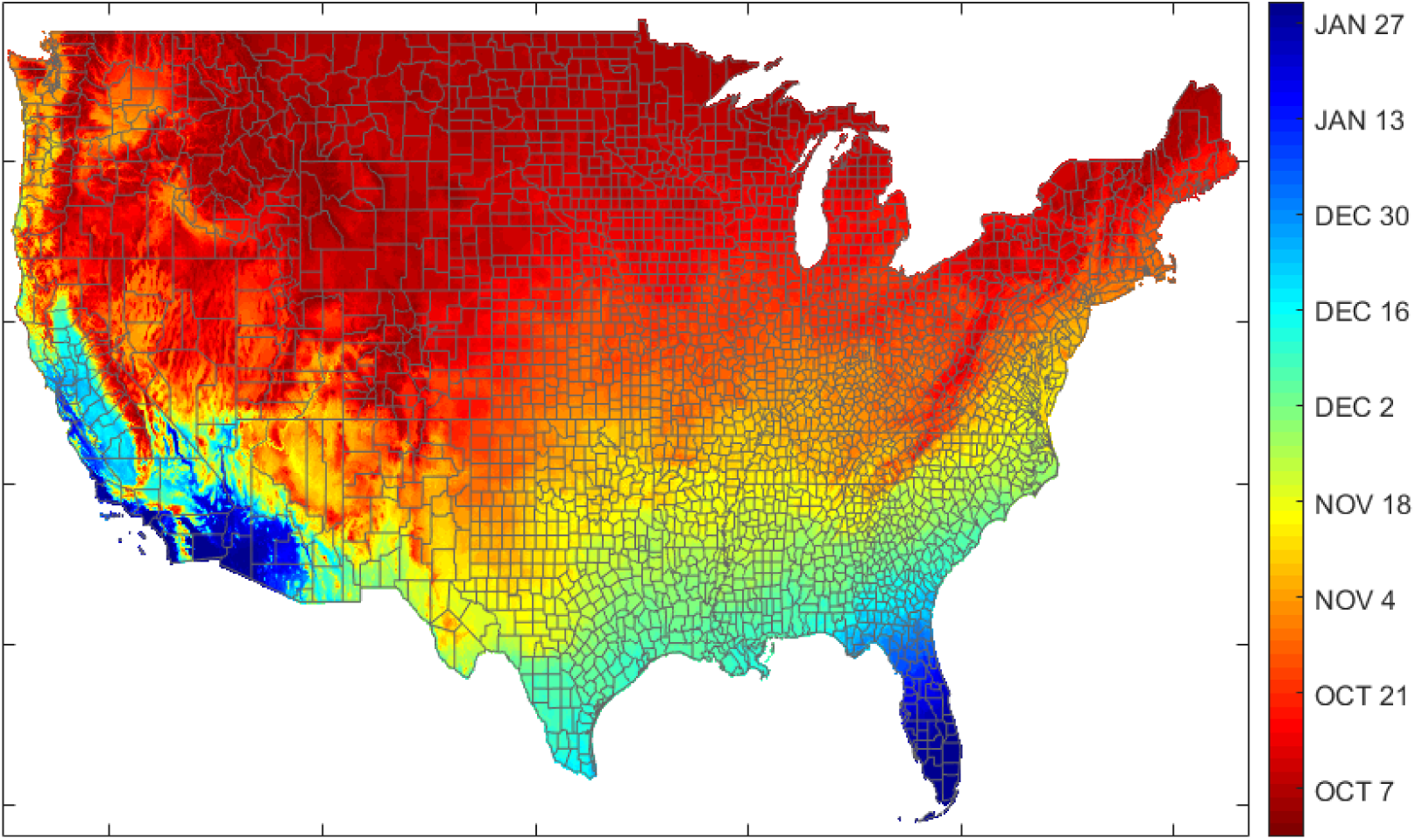
Expected date by U.S. county for entering 5-10°C range (30-year average).

There has long been fear of facing COVID-19 during influenza’s October to March season *(23)*. Our results suggest, that in addition to this concern, COVID-19 transmission will become increasingly more efficient at transmission than it was during the summer. Further, if deaths and positive case TRPs behave like influenza *(6)*, both will continue to increase from 5°C until a few degrees below freezing.

## Data Availability

All data and code from the paper will be archived at github before publication.

## Funding

The University of California, San Diego provided partial financial support for the work reported on in this paper. No outside support was received.

## Competing interests

The authors declare no competing interests. Contributions to this paper by S.L. Carson and T.K. Dye were done outside of their usual employment.

## Author contributions

**Richard T. Carson:** Conceptualization, Methodology, Software, Validation, Formal Analysis, Investigation, Data Curation, Writing – Original Draft, Visualization, Supervision, Project Administration.

**Samuel L. Carson:** Investigation, Data Curation, Writing – Review and Editing. **Thayne Keegan Dye:** Software, Investigation, Data Curation, Writing – Review and Editing, Visualization.

**Samuel A. Mayfield:** Software, Investigation, Data Curation, Writing – Review and Editing, Visualization.

**Daniel C. Moyer:** Methodology, Investigation, Writing – Review and Editing. **Chu A. (Alex) Yu:** Methodology, Formal Analysis, Investigation, Writing – Review and Editing, Visualization.

**Corresponding author biographical sketch:** Dr. Richard Carson is a Distinguished Professor of Economics at UC San Diego. His research interests focus on assessing the role of climate change and other environmental variables on a range of impacts including health outcomes.

## Appendix

### Data Preparation

The data set used in this paper starts with the COVID-19 statistics reported by individual states as aggregated daily by The COVID Tracking Project (covidtracking.com). In preliminary work, we (and other modelling groups) found there was no plausible epidemiological model that would produce the wild up-and-down jumps in the state-level COVID-19 statistics reported daily. As a result, there is a major fork in the path of any modeling effort. Do you build a predictive model for the reported COVID-19 daily death count that policymakers see, where success is judged by minimizing the forecast error around those observable quantities? Or do you attempt to model the underlying process with an eye toward understanding key aspects of it? When the reported dependent variable systematically diverges from that being generated by the underlying process, these two approaches also fundamentally diverge. For the first path, success, to some degree, comes in uncovering the administrative procedures influencing the divergence. The second path led us to undertake a major effort to rectify and repair reported COVID-19 statistics, with a focus on recovering the temporal structure needed to provide TRP estimates.

The most important correction was replacing originally reported death counts with death counts by date of death as reflected on death certificates. Figures 1 and A1 illustrate the nature of the differences for between the original CTPDailyDead_it_ and DailyDead_it_ for four important states: Arizona, Florida, Georgia and Texas. The originally reported data results in substantially larger confidence intervals in predictive models and, because it lags the revised death counts, substantially reduces one’s ability to detect and respond to changes in COVID-19 activity. We were able to make this correction for 26 states where deaths by death certificate information could be located. Our cutoff date of July 15 allowed for two and half months for states to make these death counts available, which should subsume most of the COVID-19 deaths in these states over our sample period. The states where we have been unable to obtain deaths by death certificate dates tend to be smaller and to have had relatively fewer COVID deaths. Typically, they did not face large peaks, which appear to be associated with increased testing delays. However, we have been unable to obtain this data from three large states – California, Illinois, and New York – and of the 20 observations (out of 4,567) with residuals from Eq. 1 whose absolute value is 50 or more, over half are from these three states.

The methods for collecting data backed by death certificates varied based on how the individual states chose to present them. Some states publish their counts based on death certificates in a downloadable format on their official COVID-19 website. Others publish them inside of longer reports in such a way that they can be hard to find, or present tables or graphs that need to be extracted by hand into spreadsheets. Because there are no official rules for how to publish this information, even the charts themselves vary in structure and presentation. Commonality across states is largely dependent on the specific software vendor they are using for their public-facing COVID-19 dashboards. Data collection procedures sometimes required hunting through source code to find full datasets within, or by hovering cursors over each bar in a bar chart individually over the collection window to make them display the precise value represented. In each case we were able to verify via labeling or official statements that the data was compiled using verified death certificate dates. It’s worth noting here that, while the CDC collects deaths by death certificate date (https://www.cdc.gov/nchs/nvss/vsrr/COVID19/index.htm),they only do so at the weekly level making it unusable for the sort of modeling effort we have undertaken and, relative to the data we have obtained from individual states, suffers from even longer reporting lags.

For states where deaths by death certificate date were not available, we first sought to determine if an individual state, either on their COVID-19 reporting “dashboard” or in a downloadable file, had deaths by reported date. These datasets often contain substantial corrections to what a state originally reported on a specific day. These include reports that missed covidtracking.com’s daily reporting deadline, more accurate end-of-day tallies (e.g., late reporting counties/hospitals), the correction of testing dumps that resulted in the appearance of spikes on certain days, the removal of duplicate death certificates, and the resolution of probable cases. When these differed from the COVID-19 death counts originally reported (COVIDTracking.com has a set of “snapshots” of the originally reported information) we used the updated state data. In some smaller states we believe, but have been unable to fully verify, that these corrected datasets are “close” to deaths by death certificate date.

Two of these types of corrections turn out to be particularly important. First, when a state misses a reporting deadline, this is typically recorded as there having been no COVID-19 events (new deaths positives, or tests) on that date. This causes the next day to contain the events that happened during the previous day. Second, “probable” cases have often been treated differently across states and time. State-level resolution of this issue removes an extraneous source of variation. As with the data on death certificate dates, these corrections by states of their originally reported COVID-19 statistics were obtained through a variety of sources, ranging from reading counts off interactive bar charts to downloadable CSV files.

Next, we corrected two obvious problems with the data. First, occasional, exceptionally large spikes accompanied by auxiliary information (e.g., a news/twitter release by the state’s department of health) noting that this spike was due to an accumulation of deaths over an extended time period, typically from one or more congregate living facilities, e.g., nursing homes and prisons. For these, the initial correction involved proportionately increasing death counts over the relevant period, with days where this would result in negative death counts not included in the reallocation. Second, we corrected reports of zero deaths on days surrounded by death counts that were sufficiently large that a zero-death count was highly unlikely. Such days are generally also characterized by a failure to report some other COVID-19 statistics (like new positive cases) and by an abnormally high death count on the following day (or two if it is a weekend). Our approach for these was to assume no reporting followed by backlogged reporting so that the indicated correction is to average counts across the two and, in some instances, three days.

We also corrected data in situations where a state had “corrected” an earlier report, but where an entire rectified data series from the state was not available. A typical example is a state that initially reported all deaths except for those in the state’s largest county, with the corrected version containing the death count for the whole state contained in a press/twitter release. A similar situation was when the data failed a logical consistency check by having the difference between the reported cumulative death counts on two consecutive days generate a negative daily death count. This typically occurs in states with small populations and few COVID-19 deaths, who, without comment, reduce their cumulative death count by one or two. Here we “rollback” that correction to the closest date that no longer produces a negative daily death count.

Similar corrections have been made to daily positive case counts and new daily test counts. The major difference is that very few states have made available positive case counts by day of test administration (rather than the day the test result is reported). This means that few states publish datasets that can readily subsume the positive case count data from The COVID Tracking project.

This is likely due to these types of information having different reporting standards. Specifically, a state eventually knows the recorded date of COVID-related death, but this information is subject to reporting delays due to waiting for confirming test results or autopsies. Because the date of death is on the death certificate, obtaining official death counts for all states is eventually feasible. However, while the lab doing the test knows the date of test administration, this information is often not shared with the state. States could require more complete reporting, but it is unclear whether the past is capturable. Total test counts are even messier due to the common practice – particularly in the earlier part of our sample period – of reporting all newly returned positive test results daily but reporting negative results inconsistently or in batches. Some negative test results (often by the state’s lab) were reported along with the positives, while other negatives (often those by private labs) were reported once a week. We performed rollbacks of antibody tests that, for a while, were mixed with diagnostic tests, but these are messy because information on the period over which antibody tests were included has rarely been disclosed. We have endeavored to average and prorate testing data where the nature of this practice could be reasonably inferred.

The influence of our data correction efforts can quickly be gleaned from Table A5, which describes four simple autoregressive models with a constant term and 7^th^ death count lag. The first uses the original “Reported” dataset (covidtracking.com) as both the source of the dependent variable, CTPDailyDead_it_ and the lagged death count. The second uses our corrected version as the “Revised” for the dependent variable and uses the originally Reported for the lagged regressor. The third uses CTPDailyDead_it_ as the dependent variable and the Revised for the lagged regressor. The fourth uses Revised for both. The parameter estimates for lagged deaths are similar in versions using the same lagged variable and substantively larger in the two versions using lagged revised counts. The R^2^ starts at 0.69 for the Reported/Reported model, stays roughly the same (0.68) if Revised are predicted from CTPDailyDead_it-7_, and increases somewhat further to 0.74 for the Reported/Revised combination. The Revised/Revised combination, though, has an R^2^ of 0.91, which clearly illustrates that the large gain in explanatory power in the base Eq. (1) model in Table A1 comes mainly from our data repair and rectification effort. Note that the models in Table A5 do set the massive NJ (June 25) reported death count outlier of 1877 (Revised death count is 16) to missing, since it is so large many modeling groups have either dropped it or prorate it over earlier time periods. The R^2^ of this Reported/Reported model using these observations falls to 0.42.

Temporal misalignment of NewPositives_it_ has received more attention than DailyDead_it_ because of the often-large gap in time between when a diagnostic test is administered and when the result is returned *(24)*. Indeed, this gap, coupled with different state testing regimes, has led most modeling groups to concentrate on predicting DailyDead_it_. We have made substantial repairs to NewPositives_it_ and NewTests_it_ data by reference to corrected state reports, and prorated rollbacks of initially reported antibody tests. We have been much less successful in locating information on NewPositives_it_ by date of test administration. However, temporal misalignment of the NewPositives_it-k_ is somewhat less important than for deaths than it might first appear because there is a shorter window for positive-to-positive transmission than the positive to death transition and because test results for hospital patients, the persistent high positivity pool, are typically returned quickly. Further, even very noisy testing information can be helpful in serving as controls for variation in state-level testing behavior over time.

#### Construction of Temperature, Humidity and Ultraviolet Radiation Data

Weather data for our main analysis are drawn from the National Centers for Environmental Information (NCEI) Integrated Surface Database (ISD), which report hourly temperature and humidity data for most airports in the world. For each state, weather variables are taken from the airport with the highest volume of commercial traffic, where the volume information is found in the Federal Aviation Administration’s 2018 Commercial Service Enplanements report. Our key variable of interest is daily maximum temperature (MaxTemp). We also look at measures of humidity and ultraviolet radiation. Hourly relative humidity is calculated as a function of hourly observed temperature and dewpoint temperature. Under the assumption of ideal gas behavior, we calculate hourly absolute humidity as well (details can be found at www.hatchability.com/Vaisala.pdf). We then pick the highest readings within each 24-hour period as daily MaxTemp, MaxRelativeHumidity and MaxAbsoluteHumidity. Minimum daily temperature is obtained by picking the lowest reading and the mean by averaging the hourly readings. Our measure of ultraviolet radiation is UV index, which provides a forecast of the expected risk of overexposure to UV radiation from the sun. UV index data at our representative airports was obtained from OpenWeather Ltd., which publishes daily UV index forecast calculated by the National Weather Service.

To calculate the expected date by U.S. county for entering the 5-10°C range (Figure 6), we use reanalysis data provided by PRISM Climate group (https://prism.oregonstate.edu/) which provide reliable weather data at a high spatial resolution of 4km by 4km for the contiguous U.S. We extract daily maximum temperature from 1990 to 2019 and count the average number of days it takes since Oct 1 for the daily maximum temperature to fall below 10°C.

#### Modelling Approach

There are two competing modelling approaches to predicting future COVID-19 deaths and positive cases. The first builds on a standard SEIR model; the other, a production function approach. The first has a strong epidemiological conceptualization and is clearly better in the early phase of a pandemic when data is scarce. The second is largely agnostic as to the underlying structure of transmission but requires dramatically more data to offset this flexibility. We follow the second, with (Eq. 1) using a simple production function approach. Conceptually, there is an infection pool with the seeds planted and various inputs (ranging from a state’s health care system to temperature) that influence the output: the number of deaths observed today. We primarily use past death counts as the infection pool proxy, take into account state-level fixed effects (amalgamating effects due to constant factors such as demographic characteristics, fixed resources like health care and transportation networks, and average differences in factors including climate variables and mobility), and a simple quadratic time trend. We exponentiate the right-hand-side variables to effectively impose the restriction that under the conditions we observe, expected death counts must be non-negative.

The use of a multiplicative scaling function incorporates the logic that temperature by itself cannot generate changes in DailyDead_it_. Because those dying at *t* became infected not on one day but rather over an extended period, some means of representing temperature in this setting is required. The two options, due to the strong correlation between closely adjacent MaxTemp_it-k_, are a distributed lag structure that imposes structure on a set of MaxTemp_it-k_, or parameterizing the scaling function as the product of individual scaling functions, each with a different MaxTemp_it-k_. We find two lags – the 7^th^ and 14^th^ – are sufficient and reasonably consistent with what is known about the biology of the virus and its methods of attack.

Model results are reasonably robust to small shifts in temperature lag positions, with the following two caveats. First, the 7^th^ lag is a natural one to use; many people’s lives follow a typical weekly pattern of contacts and activities, and administrative reporting procedures often have a day-of-the week pattern. Second, lags too far back are insignificant. Notably we use the 18th lag of MaxTemp_it_ in the weekly model rather than its naturally shifted counterpart LogMaxTemp_it-21_.

The most commonly used scaling function for our purposes is the logistic function 1/(1 + EXP(X)^ϒ^), where X is the variable of interest, and ϒ is the single estimated parameter. This function converges to a constant as ϒ goes to zero. Use of MaxTemp_it_ rather than LogMaxTemp_it-k_ as the stimulus variable provides a function with a different curvature. Statistically, it provides a similar fit, largely because the corresponding shifts in the estimate of ϒ makes the two scaling functions reasonably similar after normalization.

We also consider another commonly used scaling function, X/(X + Ψ), where an estimate of Ψ > 0 results in smaller values of X being scaled up more than large values of X and an estimate of Ψ < 0 results in larger values of X being scaled up more than smaller values of X, and an estimate of Ψ = 0 results in a function exhibiting no temperature responsiveness. This function can be used with either LogMaxTemp_it-k_ or MaxTemp_it-k_, and both will approximate a reasonable range of weakly monotonic scaling functions. It is well behaved as long as the estimate of -Ψ is bounded away from MIN(X), which appears not to be an issue in the situations we examine. Table A4 provides estimates for a set of competing specifications and their TRPs are displayed in Figure A8.

Our first major empirical decision was to use the three-month period of April 16-July 15. This time window allows for the seeding of the virus (to different degrees) across the i=1, …, 51 U.S. states (including DC) over a three-month period. We effectively start tracking the observable outcomes, deaths, positive cases, and tests on April 9^th^, because we generally use a one-week lag of the COVID-19 statistics of interest. Our time variable, t=1, …, 137 denoted in days starts with 1 on March 1^st^, the approximate date individual states first started reporting COVID-19 statistics.

Our second major decision involved how to implement the core epidemiological concept of a pool of infected individuals who can potentially infect other individuals. This pool is dynamic; existing positives transition to being no longer infectious, while newly infected individuals enter the pool. The totality of the currently infected individuals is unobservable without universal administration at each point in time of a 100% accurate diagnostic test. The question, thus, is whether to use a lagged variant of the test diagnosed positives, NewPositives_it_, which logically are part of the infection pool but not necessarily representative of it due to differential testing, or a lagged variant of DailyDead_it_.

The lagged DailyDead_it_ measure is one step removed but was previously assumed to always be observed *(25,26)*. This assumption is demonstrably false. In the early run of the pandemic many deaths failed to be classified as COVID-19-related, in part because some instances were thought to be influenza-related, while COVID-19’s role in inducing cardiac and kidney failure was not widely recognized. We avoid many of these problems by only using DailyDead_it_ data generated after the pandemic was well established.

Other problems with using lagged DailyDead_it_ (or NewPositives_it_) as the infection pool indicator (such as a state having a proportionately larger elderly or disadvantaged population) are readily addressed using the standard statistical approach of including time-invariant state-level fixed effects.

One way the use of a lagged version DailyDead_it_ vs. NewPositives_it_ will vary is with respect to timing. Current positives are temporally closer to future positives or deaths than current deaths and can potentially pick up the virus spreading among healthy young adults. The shorter link between a lagged NewPositives_it_ is enhanced by the short period over which a current positive can influence future COVID outcomes, which makes TRP estimation less sensitive to the choice of temperature lags.

Subject to the same measurement error, using NewPositives_it-7_ should be preferred to DailyDead_it-7_ as the infection pool indicator. These results are provided in Table A6 (Model 16). The main lesson is that the model using LogNewPositives_it-7_ is a good predictor of DailyDead_it_ (R^2^ = 0.94), but not as good as LogDailyDead_it-7_. This suggests that either the positives have more measurement error than our corrected version of death counts or that the DailyDead_it-7_ is a better reflection of the current risk-adjusted infection pool than NewPositives_it-7_. The other result worth noting is that with the infection pool indicator temporally advanced, the coefficient on LogMaxTemp_it-14_, as expected, is no longer significant.

The parameter estimates for the base model (Table A1) can be used to provide an estimate of the minimum infection pool each state faces by setting DailyDead_it-7_ = 0 (from a statistical perspective), obtaining expected DailyDead_it_, and dividing it by the CDC’s point estimate of the infection fatality rate (0.0065) *(27)*. Setting the value of Days_t_ equal to the end of the sample period, across states, the sum of these minimum infection pools is 9853 cases. Figure A3(A) provides a visual display of this information for the continental U.S. states.

#### Interpretation of State-level Fixed Effects and Time-Related Effects

We calculate a variant of the state-level fixed effects from our main regression model (Table A1) by setting the time trend equal to the last day of our sample period, the lagged number of deaths equal to zero, and calculating the expected number of deaths for each state. Small, isolated states like Hawaii generate estimates close to zero while the most populous states tend to generate estimates above 2. These estimates suggest the minimum infection pool in each state varies by a factor of approximately 20 (Figure A3(A)). A regression (Table A3) of the minimum expected death count on a small set of state-level demographics taken from the U.S. Census Bureau (LogPopulation, %Black, %Hispanic, and %Age80+) explains 79% of the variation in these counts.

We do not want our TRP estimates to be confounded by other factors changing over time (ranging from state and locally mandated shelter-in-place orders, reopening plans, endogenous social distancing, propensity to wear face masks and changing fatality rates). Since we are agnostic as to the mechanism, the straightforward specification is a polynomial time trend, where we find a quadratic term justified. Higher order terms add little insight or predictive power. The quadratic trend for predicting DailyDead_it_ falls sharply from mid-April to the end of May, after which it very slowly starts to turn up.

A model which adds (a) the number of days since a state issued a mandatory shelter-in-place order and (b) the number of days since a state started to formally reopen its economy can be found in Table A6 (Model 8). The coefficients on these two variables are small and insignificant. The two LogMaxTemp_it-k_ parameters fell on average by less than 1%.

This might seem puzzling until it is recognized that the two sets of time-related variables are highly correlated. Dropping the quadratic trend (Model 9 in Table A6) provides a different picture because, while still statistically significant at conventional levels, it was substantially diminished. The earlier a state shelter in-place order was issued, the lower the predicted death count (p = 0.028), and the earlier a state started to reopen the higher the predicted death count (p < 0.001). We are reluctant to provide any substantive interpretation to this result. Papers that have tried to determine the role of state actions and the behavior they are intended to influence show the need to (a) extensively model both state and local orders, and (b) incorporate spatially disaggregated mobility data in order to identify the effects of these government mandates from endogenous social distancing by the public that often occurs before these actions *(28,29,30)*. The two temperature coefficients in this model fall on average by less than 20% and remain highly significant.

#### Alternative Specifications for Base Death Count Model

Table A4 compares our base model to alternative specifications. These alternative specifications were chosen to look at the sensitivity of the implied TRP because they all have reasonably similar fits relative to our base model. This allows us to observe how robust the TRP is to a substantial shift in various modelling decisions that we made. The first specification replaces the LogMaxTemp_it-k_ with their linear counterparts. On the surface, this seems like a decision about which of two different scales fits better, but because the models have estimated parameters in the scale function, it may be possible for the two different specifications to provide reasonably similar TRPs. The second uses a popular ratio scaling function LogMaxTemp_it-k_/(LogMaxTemp_it-k_ + α), where α is an estimated parameter. The third replaces LogMaxTemp_it-k_ with MaxTemp_it-k_ in this ratio scaling function. A potential issue with Eq. 1, is the possibility that LogMaxTemp_it-14_ directly influences our main infection pool indicator LogDailyDead_it-7_. Our next specification replaces our infection pool indicator with an alternative, LogDailyDead_it-14_, so both temperature variables are now clearly exogenous from the temporal perspective of the infection pool indicator.

The last is a weekly variant (Eq. 2) of the base model, where the dependent variable is the sum of the daily death count of the next seven days with corresponding pushbacks of the lagged variables. This model averages out much of the daily variation and many types of administrative reporting practices. In forecasting the pandemic’s progression, it has become common to use data aggregated to a weekly level in an effort to average out many of the administratively-induced reporting issues that our extensive reconstruction and repair of the daily death count data sought to alleviate. It is possible to estimate a variant of Eq. (1) that uses death count data aggregated into seven-day periods, WeeklyDead_it_ = Σ_t_ DailyDead_it_, where the summation is over t=1 to t=7. This makes WeeklyDead_it-7_ the sum of the 7th through 13th lags of DailyDead_it_. Importantly, this aggregation does not reduce the number of observations because on each day, the weekly aggregation at t=1 adds a new observation DailyDead_i1_ and drops DailyDead_i8_. Lags of the WeeklyDead_it_ variable can then be used in the standard way. One implication of this specification is that the temperature variables also need to shift backwards. For conceptual consistency, we use LogMaxTemp_it-14_ in place of LogMaxTemp_it-7_. Empirically, the model fits best with the second temperature lag being LogMaxTemp_t-18_. The model fit is reasonably similar using the 13th lag and 19th lags, but beyond the 19th lag, the temperature variable becomes insignificant, suggesting temperature information farther back in time than this is not useful. The model we report is thus:

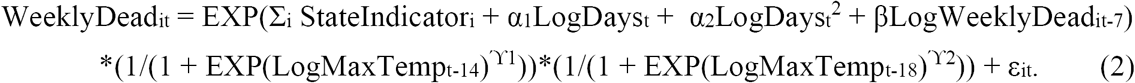

The overall impression from Table A4 is the general stability of most of the common parameters for time and the infection pool. Some of the specifications offer insights into the latter variable. It is little influenced by whether the logistic or ratio scaling function was used, nor whether LogMaxTemp_it_ or MaxTemp_it_ was the stimulus variable. Using LogDailyDead_it-14_ in place of LogDailyDead_it-7_ results in an estimated TRP being similar to the base model (Figure A6), although it does rise more sharply between 10°C and 5°C suggesting, while there may be a endogeneity effect, it is not large and that our base TRP is conservative. In this model (Table A6, Model 7), the coefficient on the infection pool indicator shifts from the .86 of the base model to .80; in the weekly version model it jumps to .92, The R^2^ for the weekly model increases to 0.99 (from the base model’s 0.97) and falls to 0.94 in the model using LogDailyDead_it-14_, which is less informative than LogDailyDead_it-7_ in term of the infection pool influencing current death counts .

#### Constructing Temperature Response Profiles (TRPs)

To compare the TRP’s implied by the models in Table A4, we plot the functions using two independent random uniform variables RTemp and RTemp2, defined over the range 5°C and 40°C. The logistic scaling function with two temperature variables, LogMaxTemp_it-7_ and LogMaxTemp_it-14_, and corresponding Eq. 1 estimated parameters ϒ1 and ϒ2 (Table A4), results in the following scaling function in the base model:

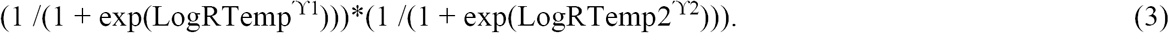

There is a fundamental indeterminacy in such a scaling function, in that multiplying the production function part of Eq. 1 by a constant will result in an offsetting change in the scaling function which maintains the same expected value for the dependent variable. Note that each of the two multiplicative components of Eq. 3 converges to .5 irrespective of temperature values as ϒ1 and ϒ2 become increasingly negative. Often logistic functions are normalized to lie between 0 and 1 by changing the “1” in the numerator to “2”, but this is not needed with our normalization to 31°C, which solves the indeterminacy from the perspective of comparing curves. This is done by calculating the value of the estimated scale function at 31°C:

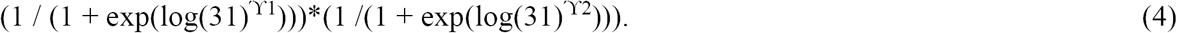

Dividing Eq. 3 by Eq. 4 produces a function which equals 1 at 31°C. Multiplying this quantity by 100 produces a function which has a natural percentage interpretation and equals 100 at 31°C. Note that for forecasting purposes, the original scaling function parameters need to be used.

The ratio scaling function with estimated parameters α_1_ and α_2_ using RTemp and RTemp2 is:

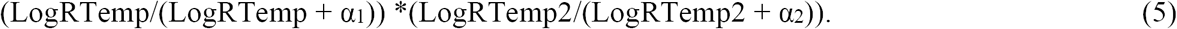

As α_1_ and α_2_ converge to zero, both multiplicative components of SI.1c converge to 1 irrespective of temperature values. The value of this function at 31°C can be calculated in a manner similar to that described for the logistic. Dividing Eq. 5 with the value of that function at 31°C and multiplying by 100 produces the desired TRP.

Figure A8 graphs the daily death TRPs from the model specifications in Table A4. The alternative specifications produce TRPs remarkably similar to that of our base model and tend to bracket it. The one systematic difference is that TRPs based on the ratio scaling functions tend to be flatter over 10°C to 20°C but rise more sharply in the 5°C to 10°C range.

#### Further DailyDead_it_ Specifications Involving Weather, Positives and Cumulative Dead

The other weather variables which have received considerable attention are absolute humidity, relative humidity, and ultraviolet (UV) radiation. Details of construction are provided below in Data Preparation Section. The limitation with all these variables is that they are correlated with MaxTemp_it_ (i.e., absolute humidity: 0.58, relative humidity: -0.21, UV: 0.79). Maximum absolute humidity has a reasonable size effect (p < 0.001) in the model where it replaces the corresponding MaxTemp_it_ variables. However, the LogAbsoluteHumidity_it-k_ parameter estimates are close to zero and are no longer significant when the two parallel logistic functions comprising the temperature scaling function are added (Model 10 in Table A6). Relative humidity has a marginally significant relationship with DailyDead_it_ when MaxTemp_it_ is not in the model and its effect is close to zero in a model with MaxTemp_it_ (Model 11 in Table A6).

The situation with UV_it_ for which (15) found support is more complex (Table A4 &Table A6, Models 12 and 13). The MaxTemp_it_ lags are marginally better predictors in a head-to-head comparison. These two variables are strongly correlated. Adding UV_it_ marginally decreases prediction errors, but the signs of high multicollinearity are obvious; the first MaxTemp_it_ lag is still highly significant while the second is insignificant and one of the UV_it_ lags is only marginally significant (Table A4). It is effectively impossible to disentangle the influence of LogMaxTemp_it_ and UV_it_. Their relationship is displayed in Figure A5, which plots LogMaxTemp_it_ and LogUV_it_ for two states over our sample period: Georgia (Atlanta) and New York (New York City). We cast our results in terms of MaxTemp_it_ rather than UV_it_ because it is more widely reported and understood, without making any claim our work supports a joint versus singular causal mechanism.

The specification in Eq. (1) is cast in terms of DailyDead_it_. If MaxTemp_it-k_ influences COVID transmission via the link between positive cases, then we should be able to replace DailyDead_it-7_ with NewPositives_it-7_. Estimates for this model (Table A4) show LogNewPositives_it-7_ being significant at p < 0.001 and the R^2^ measure falling a bit. The coefficient on MaxTemp_it-14_ is insignificant, which would be expected if NewPositives_it-7_ incorporates that information, while the coefficient on LogMaxTemp_it-7_ is larger than in the base specification.

A different aspect of the COVID-19 death statistics that has not been incorporated into the model is lagged cumulative death count, TotalDead_it-k._ If DailyDead_it-k_ can be seen as a proxy for the infection pool influencing DailyDead_it_, then TotalDead_it-k_ (normalized by population) is proxy for the fraction of the population that no longer at risk in the sense of being either removed by death or recovered. Adding LogPCTotalDead_it-7_ effectively makes the StateBase_it_ dynamic in a potentially different way than the quadratic time trend by letting each state evolve according to its own pattern of deaths. Table A6 (Models 14 and 15) displays the results of this model with LogPCTotalDead_it-7_ enter as (a) a second order polynomial and (b) a fourth order polynomial.

Three results are worth noting. First, the increase in explanatory power is small with the most noticeable changes being as expected in the StateIndicator_i_ and a substantial reduction in importance of the overall quadratic time trend. Second, the LogMaxTemp_it-k_ parameter estimates are similar to that of Eq. (1) suggesting that our TRP is robust to a substantial dynamic reparameterization of the model. Third, in the quadratic specification, DailyDead_it_ is declining with as LogPCTotalDead_it-7_ at a declining rate. In the fourth order specification, all the LogPCTotalDead_it-7_ terms are insignificant (although jointly significant). Figure A6 displays, starting at .05, the two response functions for LogPCTotalDead_it-7_. Like the quadratic time trend, they suggest a sharp drop in how DailyDead_it_ is influence by DailyDead_it-7_ as LogPCTotalDead_it-7_ increases from low levels with the fourth order polynomial being flatter at high levels of LogPCTotalDead_it-7_ than the quadratic. The influence of this factor is reasonably small by the time a state hits 10 deaths per 100,000, a condition that characterizes 80% of the states at the end of our sample period. Earlier, we noted one interpretation of the quadratic time trend was that medical care (and hence death rates) had improved sharply at first and then at a declining rate. This specification has a similar interpretation but suggests that some of that learning is state specific and related to its prior COVID-19 caseload. There is no indication that this rate of decline is accelerating even in the hard-hit Northeastern states where deaths per 100,000 can be as high as 175 (NJ), looking at the fourth order polynomial which should allow this feature to emerge if the data supports it. This suggests the magnitude of the fraction of the population previously infection is still too small to be a substantial factor in slowing transmission of the virus.

#### Univariate DailyDead_it_ and MaxTemp_it-7_ Relationship

If we have succeeded in isolating the TRP through use of the set of StateIndicator_i_ and a quadratic time trend, the simple regression of DailyDead_it_ on lagged MaxTemp_it-7_ without the state fixed effects and quadratic time trend should reveal a substantially different curve than our estimated TRP displayed in Figure 4. Figure A7 displays this relationship using a robust LOWESS smoother (bandwidth 0.2) on MaxTemp_it-7_. This curve is dramatically more sensitive to temperature between 10°C and 30°C. Below 10°C it drops, which was expected since states with temperatures near 5°C tend to be more isolated and smaller population-wise. The curve bends up near 35°C and beyond where most of the observations come from a few states with (relatively) high June and July death counts.

Pinning down the TRP further will require: (a) obtaining more data over a longer time horizon with more temperature variation and, in particular, the important -5°C to 5°C range, where there has been little U.S. experience since the virus became widely dispersed, (b) obtaining death certificate data information from the few remaining large states (California, Illinois and New York) where it is not yet available, since states with death certificate date reporting have prediction errors that are substantially smaller (p < 0.001) than those that don’t (Table A6, Model 2), or (c) having high quality temporally aligned COVID-19 statistics at the county level, which would provide a better temperature match and dramatically increase sample size.

#### Death Count TRPs based on DailyDead vs. COVID Tracking Project Originally Reported Counts

What does a TRP based on The COVID Tracking Project’s (CTP) originally reported death counts look like compared to our base (Eq. 1) Model 1 which uses DailyDead_it_? To examine this issue, we estimate Model 26 (Table A6), a direct analogue of Model 1, that substitutes CTPDailyDead_it_ and CTPDailyDead_it-7_ for their DailyDead_it_ counterparts. Consistent with Model 18 (Table A6) and other models employing CTPDailyDead_it_, we exclude the two observations that contain the large New Jersey outlier as either the dependent variable or regressor and further excluded twenty observations where LogCTPDailyDead_it-7_ is undefined because CTPDailyDead_it-7_ is negative.

There are some clear differences between Model 1 and Model 26. First, in Model 26 there is a large drop in the R^2^ from 0.97 to 0.81 and the RMSE measure more than doubles. Second, the elasticity parameter on lagged LogCTPDailyDead_it_ is just over 60% as large as it is in the DailyDead_it_ version. This is the expected result from introducing substantial measurement error, but one that also has large implications for drawing inferences about the size of the infection pool or in undertaking any dynamic forecasting exercise. Third, the quadratic time trend, while still sizeable, is substantially diminished in both magnitude and statistical significance. Fourth, there are differences between the TRPs implied by their temperature response parameters in Eq. (1).

The TRPs for Model 1 and Model 26 are plotted in Figure A9. The TRPs for the CTPDailyDead_it_ variant effectively lies above its DailyDead_it_ counterpart. At 5°C, it predicts almost 80% more deaths than our base Model 1. This result may seem counterintuitive to the usual belief that measurement error induces the parameter estimate to be attenuated toward zero. That, intuition, generally correct, helps explain the fall in coefficient when CTPDailyDead_it-7_ rather than DailyDead_it-7_ is used as the infection pool indicator. However, it does not say anything about the implication of introducing measurement error into another covariate in the model. It is easy to show that as the scale of the measurement error in the lagged death count increases, the magnitude of the temperature responsiveness parameters adjusts to incorporate covariance with DailyDead_it-k_. All else held constant, this would increase the magnitude of the estimated temperature effect under classical measurement error, since the parameters on infection pool indicator and temperature variables should have the same sign.

The situation here is more complicated because measurement error from using CTPDailyDead_it_ also substantially reduces the ability to pin down the quadratic time trend. Thus, neither the sign nor magnitude of the difference between the temperature response parameters across comparable specifications is therefore known a priori. Our parameter estimates (Table A6) suggest an upward bias is likely in temperature effect estimates obtained from models similar to ours that (a) use reported death counts and (b) have specifications where the infection pool and temperature response variables are expected to share the same sign. Note though that in a simple OLS regression model, the infection pool and temperature variables should have opposite signs, resulting in an estimate of the temperature parameter likely biased in the direction of finding no effect.

#### Structure of Table A6

The parameter estimates and standard summary statistics for models discussed is this paper are provided in Table A6. It contains three columns for each model. The first column contains the variables included in the model, the second the parameter estimates, and the third the standard errors. After the parameter estimates, the model’s R^2^, root mean square error (RMSE), and the number of observations on which the model was fit are provided.

The order in which the models appear are:

1. The base model represented by Eq. 1 using LogDays_t_, LogDays_t_^2^, and LogDailyDead_it-7_ as the predictors along with a set of state-level indicator variables and using LogMaxTemp_it-7_ and LogMaxTemp_it-14_ in the temperature scaling function.
2. A model which regresses the squared residuals from (1) on the log of state population and an indicator variable, GOOD_DATE for DailyDead_it_ representing death counts by death certificate date.
3. Base model using linear versions of the two temperature variables.
4. Base model using alternative ratio scaling function Temp/(Temp + α), where Temp is temperature variable and α is the estimated parameter.
5. Base model using the alternative ratio scaling function and linear versions of the MaxTemp_it-k_.
6. Base model with LogDailyDead_it-14_ substituted for LogDailyDead_it-7_.
7. A weekly version of the base model (see Eq. 2) substituting LogDailyDead_it-14_ for LogDailyDead_it-7_ and LogDailyDead_it-18_ for LogDailyDead_it-14_.
8. A version of the base model that adds two state-level government policy variables, the log of the number of days since a shelter-in-place order was first issued (LogDaysShelterInPlace_it_) and the log of the number of days since a state began to formally reopen its economy (LogDaysReopen_it_).
9. A version of (8) that drops the quadratic time trend of the base model.
10. The base model adding parallel scaling functions using the LogMaxAbsoluteHumidity_it-7_ and LogMaxAbsoluteHumidity_it-14_.
11. The base model adding parallel scaling functions LogMaxRelativeHumidity_it-7_ and LogMaxRelativeHumidity_it-7_.
12. Base model adding additional parallel scaling functions for LogUV_it-7_ and LogUV_it-14_.
13. Base model substituting LogUV_it-k_ for LogMaxTemp_it-k_.
14. Base modeling adding a quadratic in terms of LogTotalDead_it-7_.
15. Base modeling adding a 4th order polynomial LogTotalDead_it-7_.
16. Base model substituting LogNewPositives_it-7_ and testing variables for LogDailyDead_it-7_.
17. The model used in the paper to predict NewPositives_it_.
18. The AR(7) regression of the originally reported death counts from The COVID Tracking Project (CPTDailyDead_it_) on the 7^th^ lag of itself (CPTDailyDead_it-7_) for Table A5.
19. The same as (18) but with DailyDead_it_ regressed on the 7^th^ lag of the CTPDailyDead_it_.
20. The same as (18) but CTPDailyDead_it_ regressed on the 7^th^ lag of DailyDead_it_.
21. The same as (18) but DailyDead_it_ regressed on its 7^th^ lag.
22. The StateBase_it_ calculation for each state.
23. The model reported in Table A3 which predicts the state-level fixed-effect estimates obtained in (1) as a function of a small set of demographic variables.
24. State-level estimates of the minimum infection pool.
25. State-level change in expected DailyDead_it_ when DailyDead_it-7_ shifts from 0 to 1 on July 15, assuming 31°C.
26. Base model (1) using the uncorrected death count data.

### Data and Code Availability

The data used in this study are archived at https://github.com/xxx in the form of a Stata “.dta” file. An Excel version of this file was created using StatTransfer. The Stata “do” file creating the data set contains a line-by-line set of the changes made to the original CovidTracking.com data set, and the providence of those changes. Three additional Stata “do” files are available in this archive. The first contains the code (Stata 16.1) for the regression models reported in this paper. The second provides an example of how to estimate the static and dynamic temperature response profiles for an individual state using Georgia as an example. The third contains Stata code for creating the basic versions (fine labeling was done using Stata’s graph editor) of the figures in this paper. We also provide a further Stata dataset and corresponding Excel files at the state level (using our representative airport) with maximum daily temperature readings from the last thirty years.

### Appendix Figures

**Figure A1.**
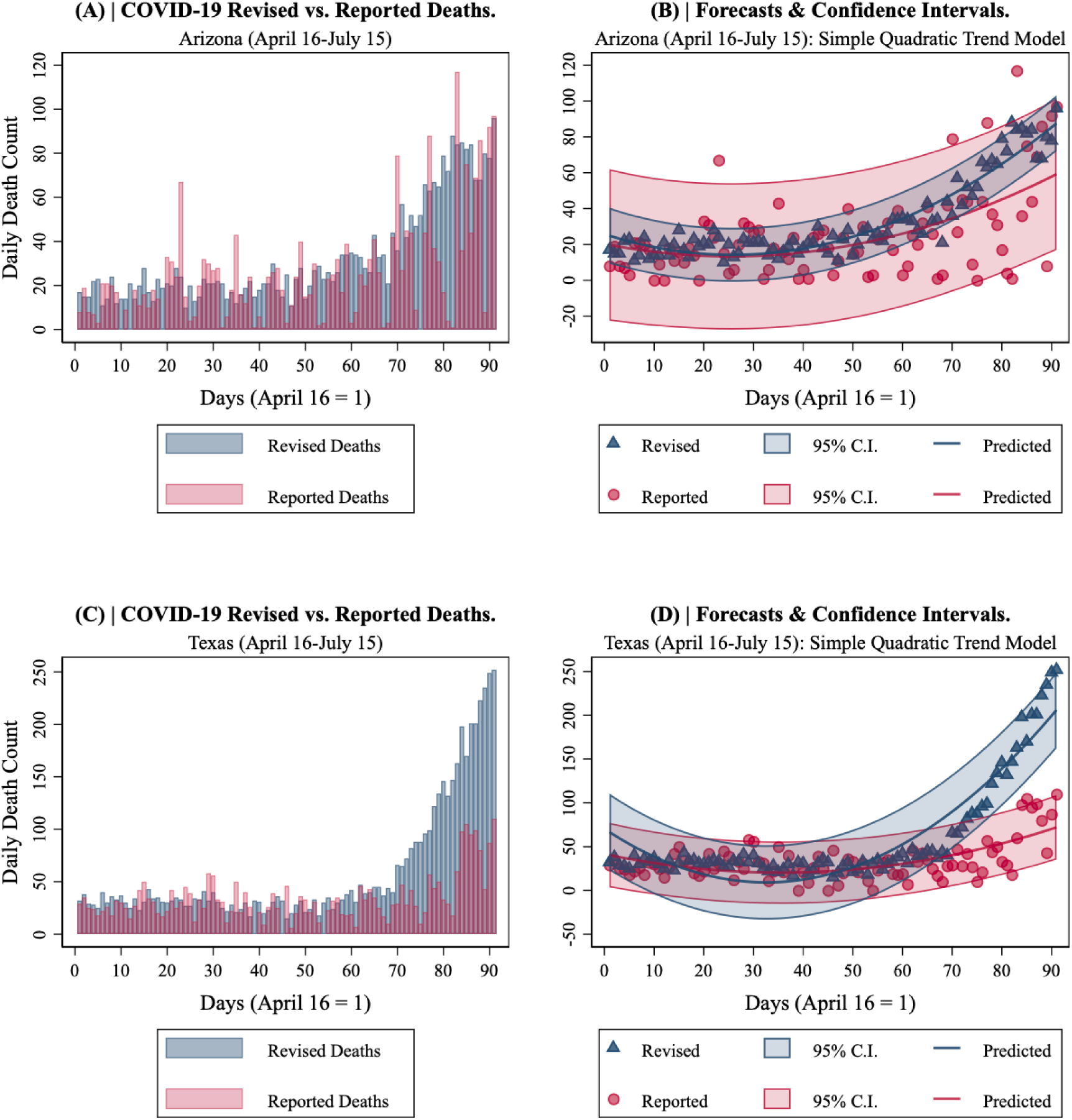
COVID-19 deaths in Arizona and Texas. April 16-July 15. **(A)** Arizona Revised vs. Reported. **(B)** Arizona quadratic time trend forecasts and confidence intervals. **(C)** Texas Revised vs. Reported. and **(D)** Texas quadratic time trend forecasts and confidence intervals.

**Figure A2.**
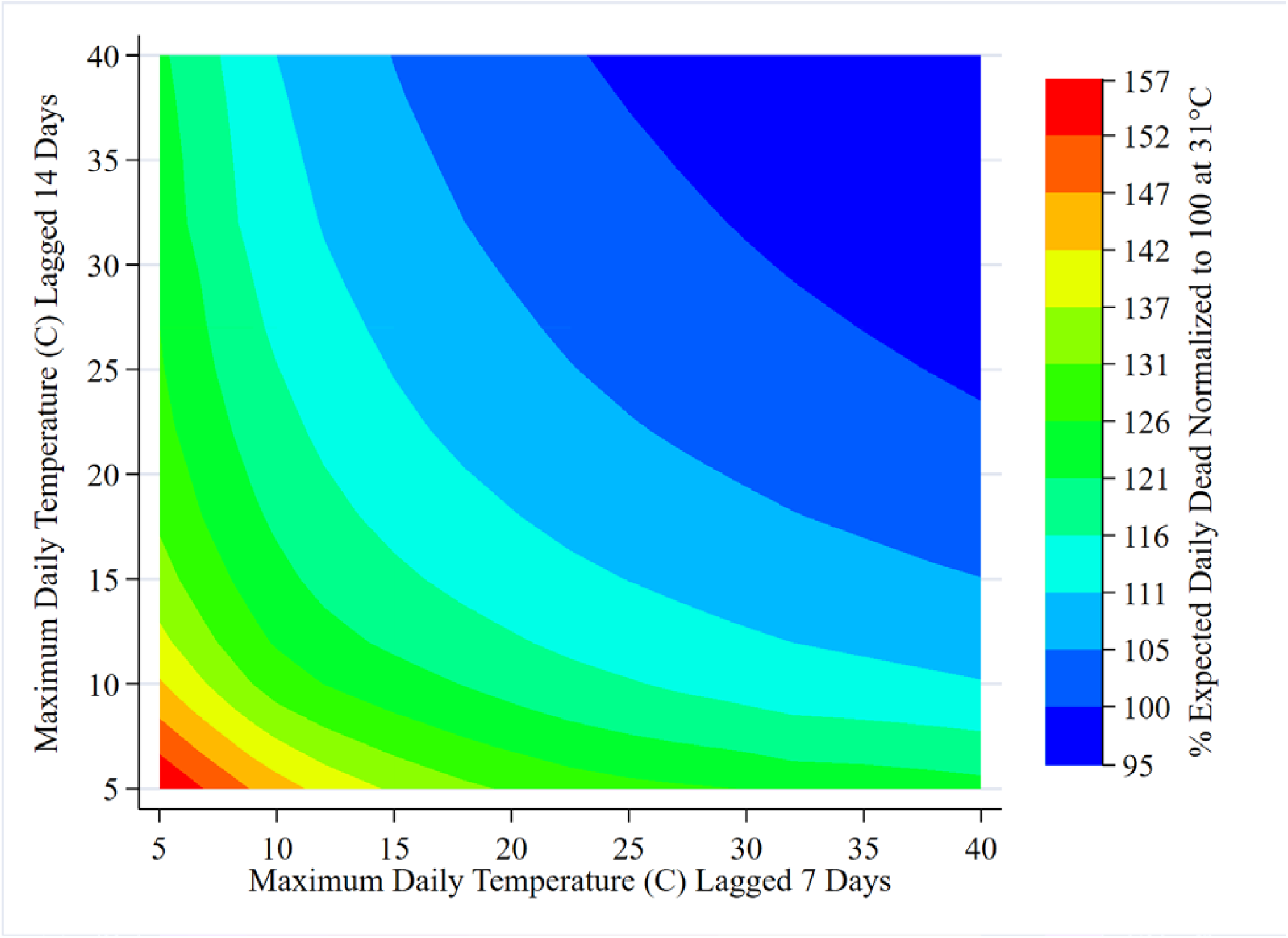
DailyDead TRP contour plot representation.

**Figure A3.**
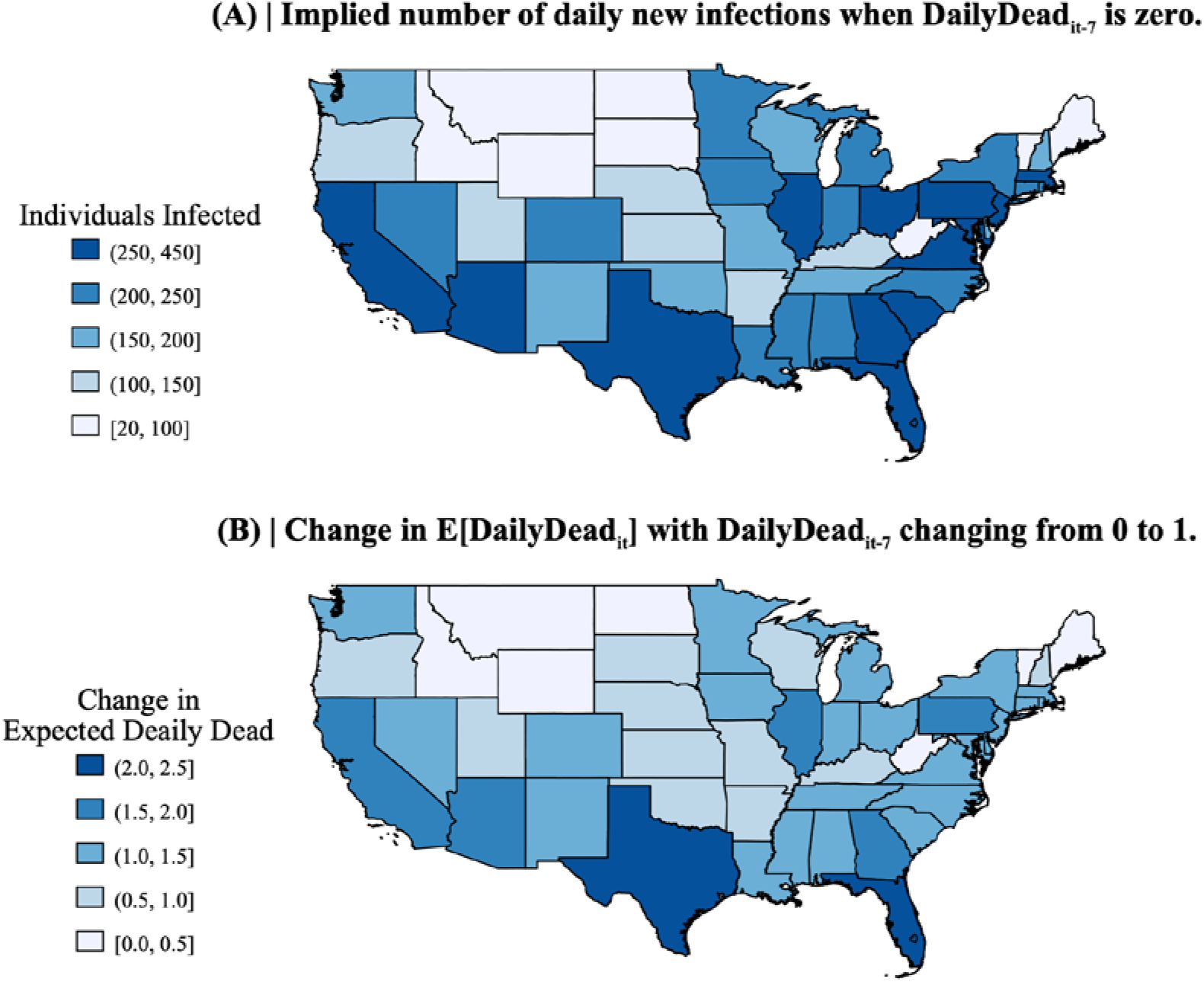
Visual representation of information contained in the parameter estimates for base model (Eq. 1). **(A)** displays the infection pool for each state in the continental U.S. implied if observed DailyDead_it-7_ is set to zero on July 15 and the two lagged MaxTemp_it-k_ are set to 31°C. **(B)** shows, under the same conditions, how the expected DailyDead_it_ changes if DailyDead_it-7_ moves from 0 to 1.

**Figure A4.**
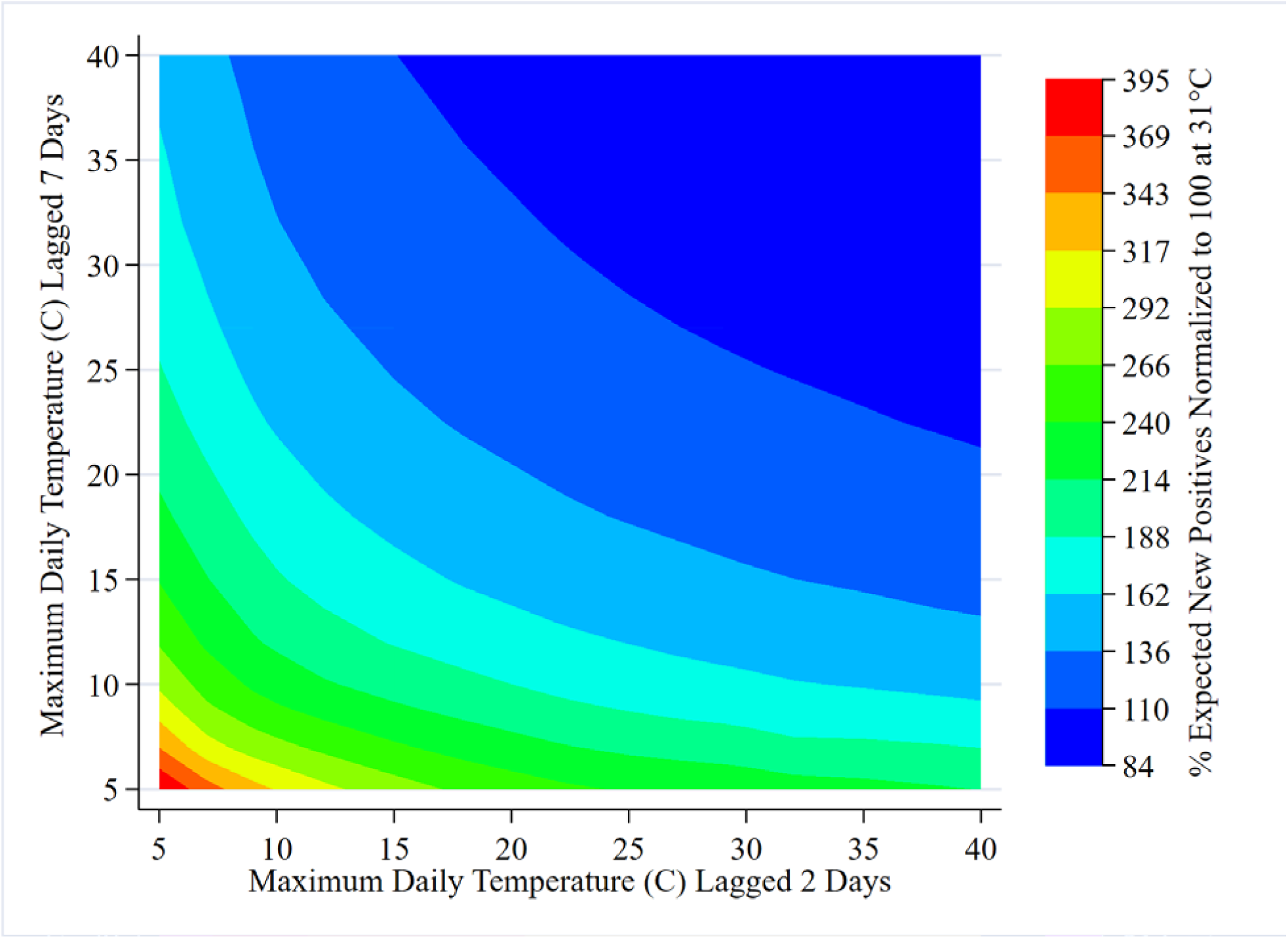
NewPositives TRP contour plot representation.

**Figure A5.**
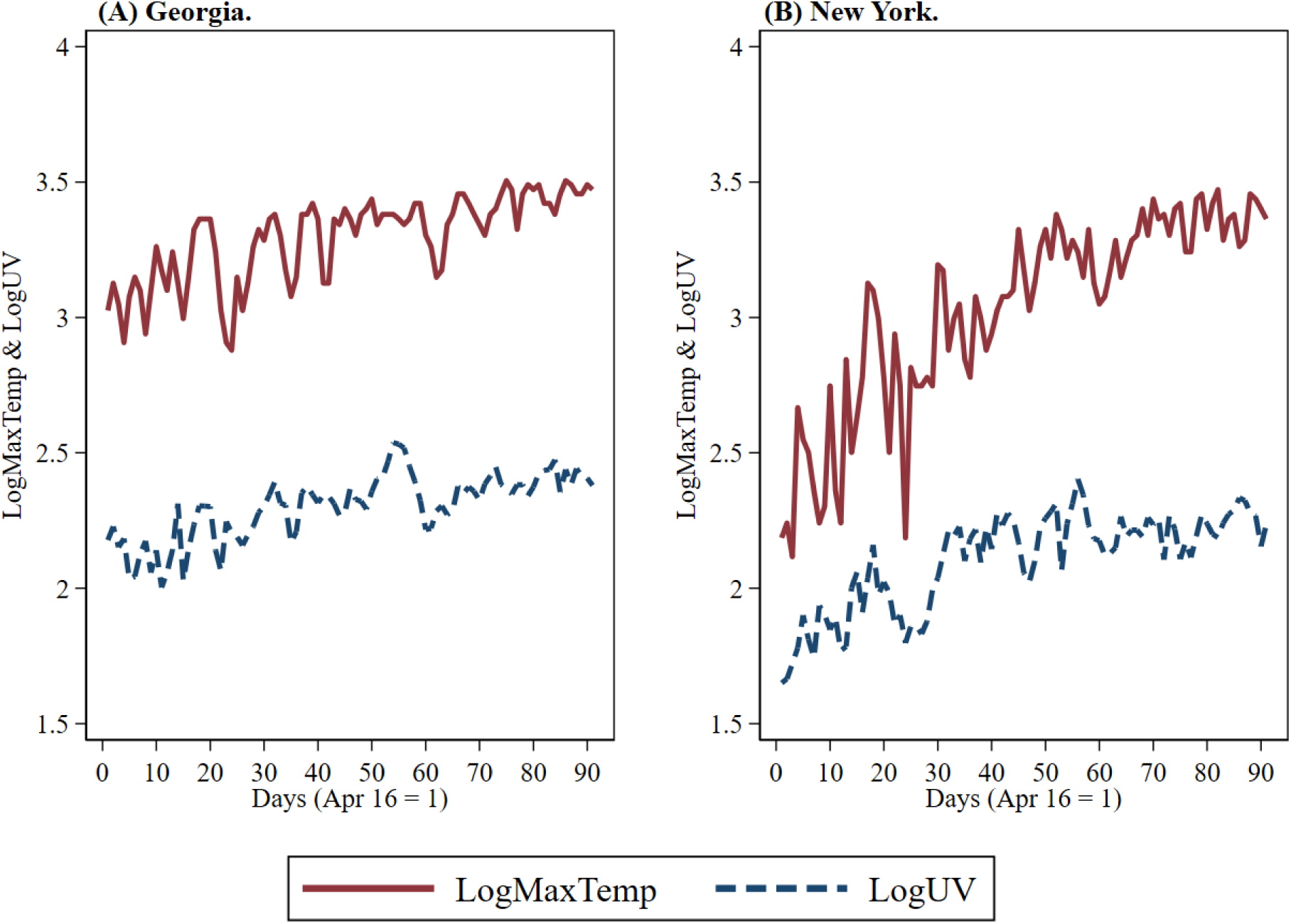
LogMaxTemp vs. LogUV. **(A)** displays Georgia. **(B)** displays New York.

**Figure A6.**
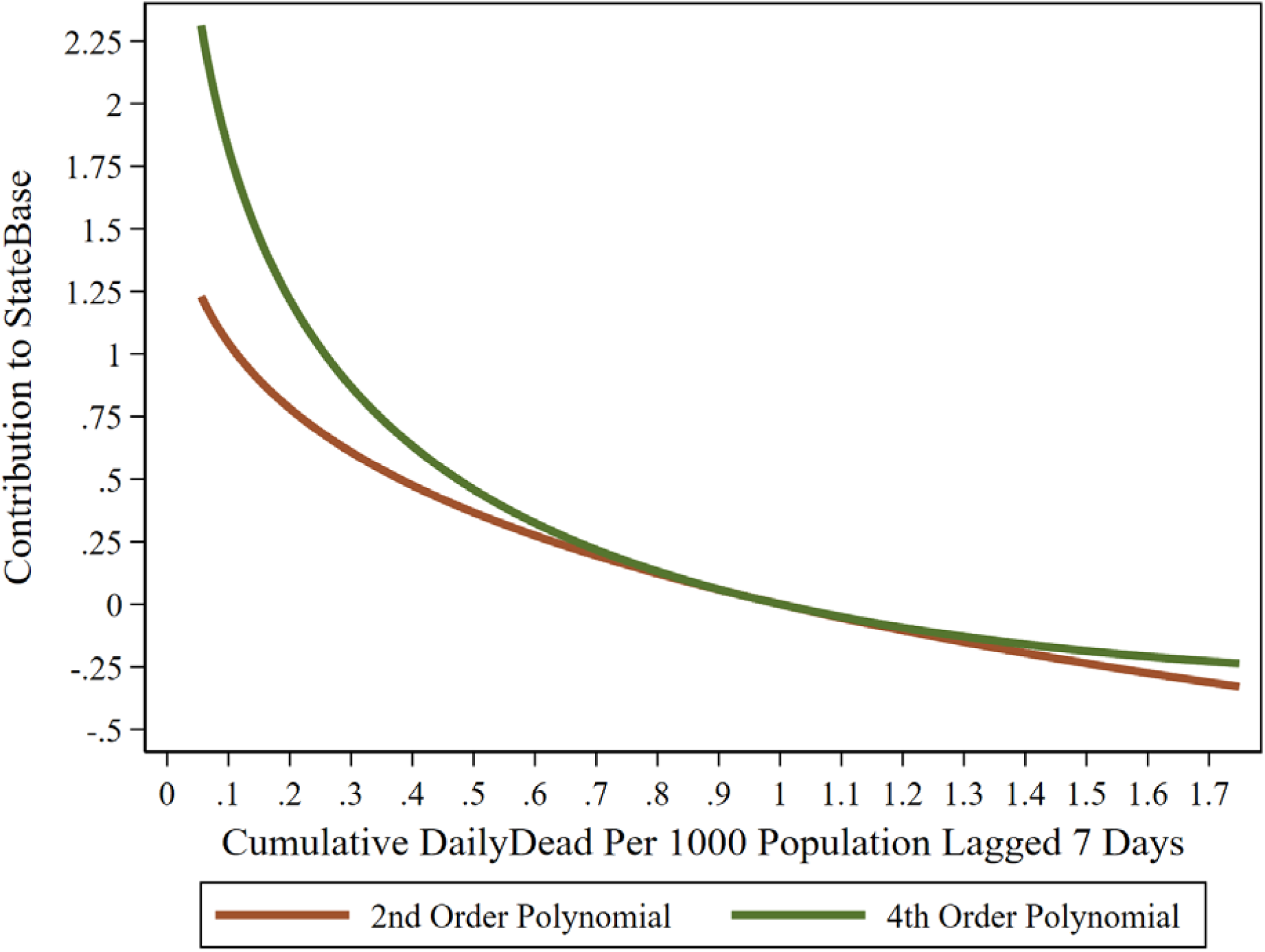
DailyDead_it_ responsiveness to lagged per capita total dead.

**Figure A7.**
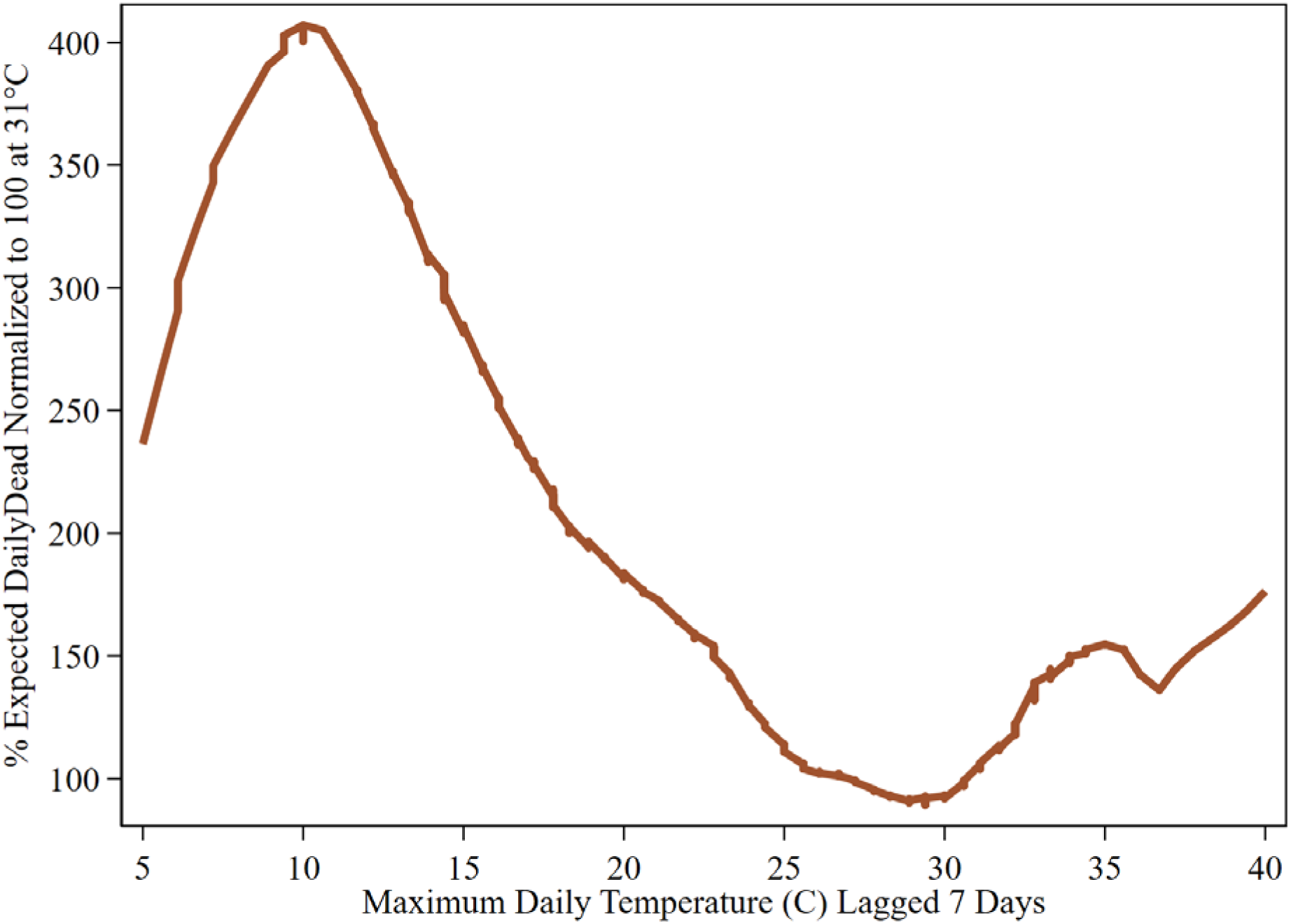
Bivariate relationship DailyDead & MaxTemp. U.S. states: April 16-July 15. Lowess smoother: .2 bandwidth.

**Figure A8.**
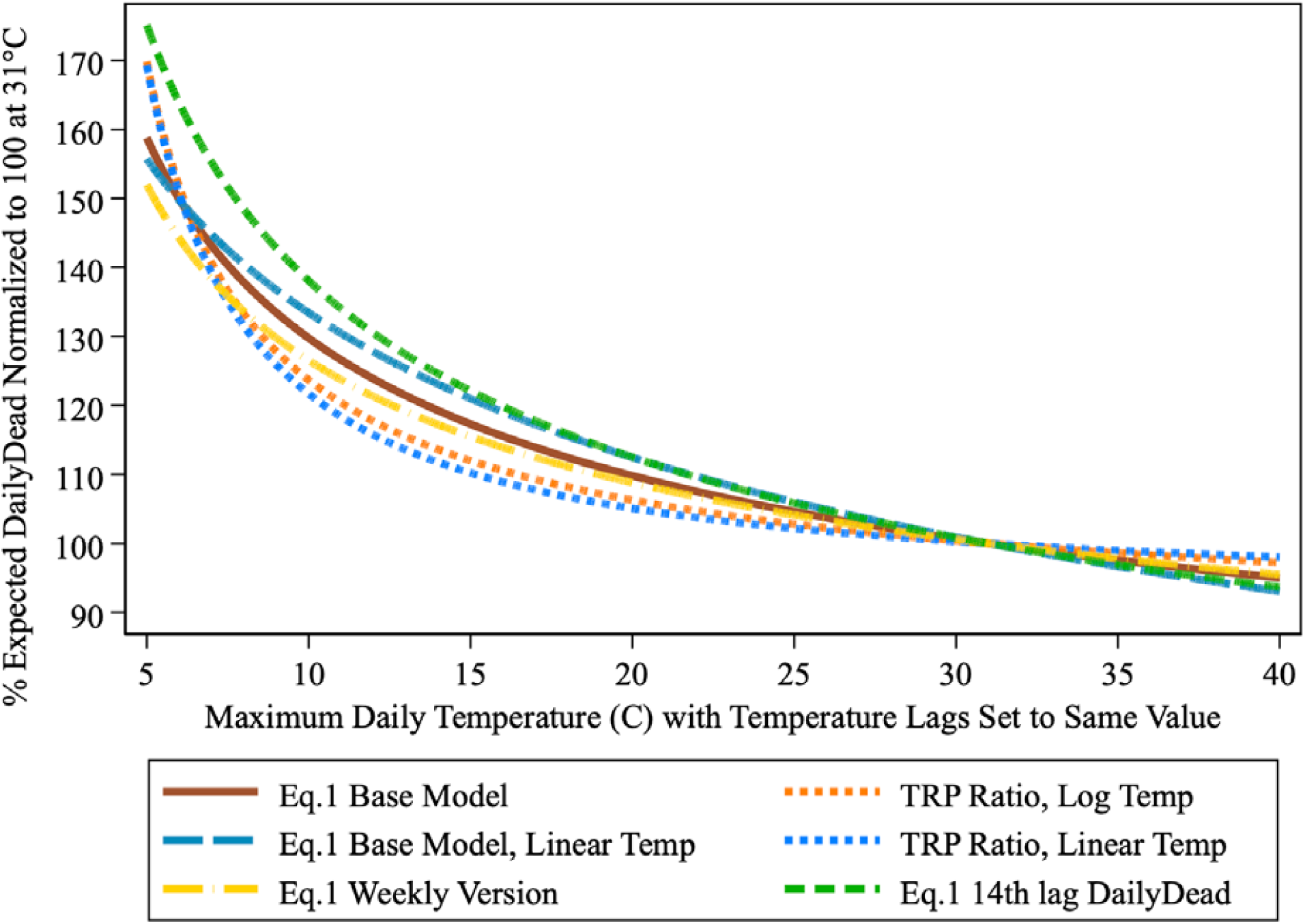
Alternative DailyDead TRPs. U.S. states: April 16-July 15.

**Figure A9.**
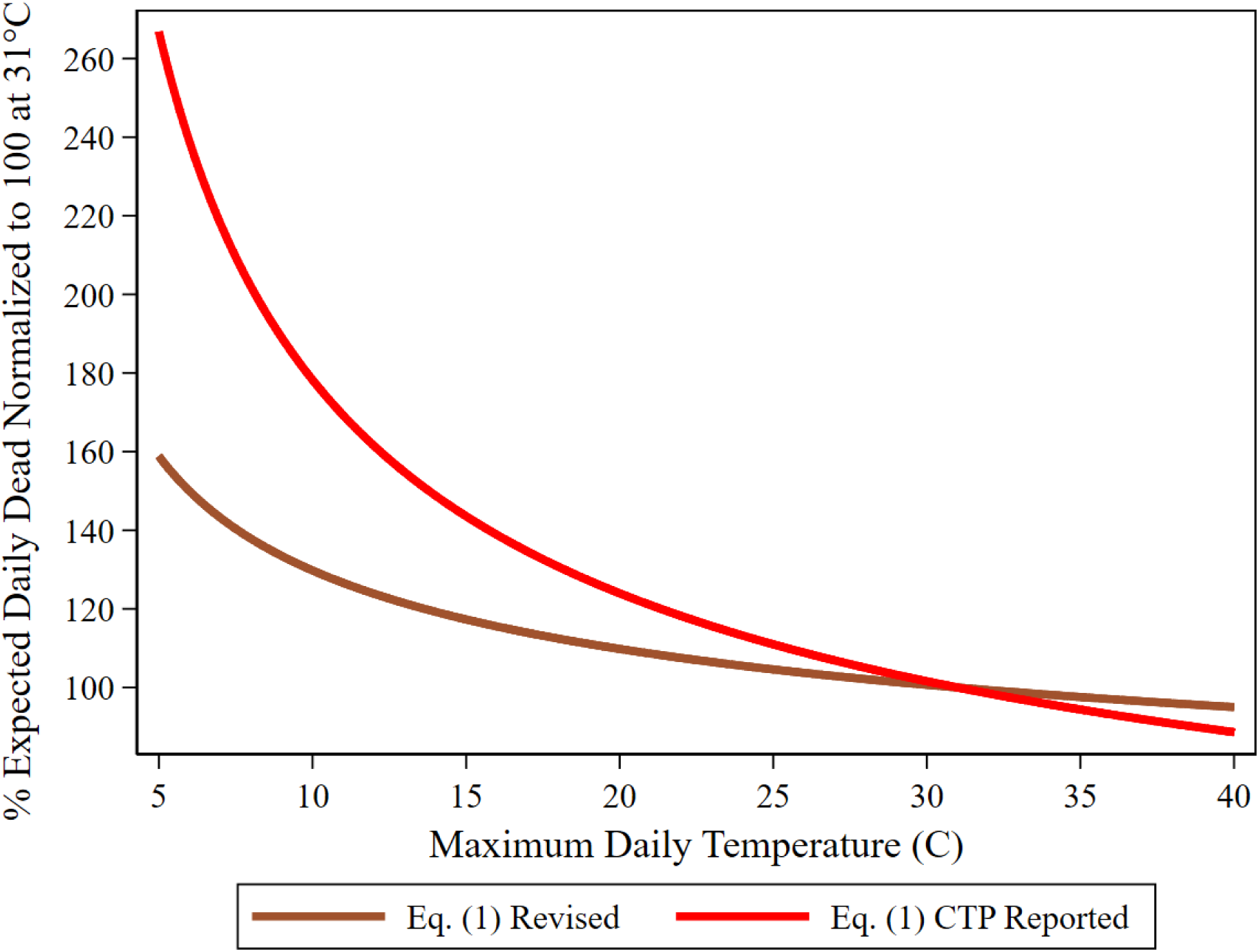
Death count TRPs based on DailyDead vs. CTP originally reported. U.S. states: April 16-July 15.

**Table A1.** Eq. (1) base model predicting DailyDead_it_.

| **Variable** | **Coefficient** | **Robust S.E.** | **t-statistic** | **p-value** |
| --- | --- | --- | --- | --- |
| Alaska | 24.2468 | 3.2668 | 7.42 | 0.000 |
| Alabama | 26.2830 | 3.2504 | 8.09 | 0.000 |
| Arkansas | 25.6412 | 3.2496 | 7.89 | 0.000 |
| Arizona | 26.6339 | 3.2277 | 8.25 | 0.000 |
| California | 26.5480 | 3.2434 | 8.19 | 0.000 |
| Colorado | 26.1724 | 3.2608 | 8.03 | 0.000 |
| Connecticut | 25.3270 | 3.2578 | 8.08 | 0.000 |
| District of Columbia | 26.0314 | 3.2699 | 7.96 | 0.000 |
| Delaware | 26.0525 | 3.2719 | 7.96 | 0.000 |
| Florida | 26.7896 | 3.2194 | 8.32 | 0.000 |
| Georgia | 26.4536 | 3.2545 | 8.13 | 0.000 |
| Hawaii | 23.8933 | 3.2676 | 7.31 | 0.000 |
| Iowa | 26.1666 | 3.2669 | 8.01 | 0.000 |
| Idaho | 25.3286 | 3.2630 | 7.76 | 0.000 |
| Illinois | 26.5186 | 3.2569 | 8.14 | 0.000 |
| Indiana | 26.3159 | 3.2619 | 8.07 | 0.000 |
| Kansas | 25.6968 | 3.2630 | 7.88 | 0.000 |
| Kentucky | 25.8271 | 3.2613 | 7.92 | 0.000 |
| Louisiana | 26.3165 | 3.2548 | 8.09 | 0.000 |
| Massachusetts | 26.4245 | 3.2557 | 8.12 | 0.000 |
| Maryland | 26.4151 | 3.2614 | 8.10 | 0.000 |
| Maine | 25.0056 | 3.2684 | 7.65 | 0.000 |
| Michigan | 26.2274 | 3.2515 | 8.07 | 0.000 |
| Minnesota | 26.3019 | 3.2628 | 8.06 | 0.000 |
| Missouri | 25.9973 | 3.2607 | 7.97 | 0.000 |
| Mississippi | 26.2978 | 3.2530 | 8.08 | 0.000 |
| Montana | 24.7679 | 3.2679 | 7.58 | 0.000 |
| North Carolina | 26.3227 | 3.2556 | 8.09 | 0.000 |
| North Dakota | 25.2256 | 3.2686 | 7.89 | 0.000 |
| Nebraska | 25.7297 | 3.2615 | 7.92 | 0.000 |
| New Hampshire | 25.8414 | 3.2635 | 7.92 | 0.000 |
| New Jersey | 26.3998 | 3.2477 | 8.13 | 0.000 |
| New Mexico | 26.1024 | 3.2650 | 7.99 | 0.000 |
| Nevada | 26.1382 | 3.2566 | 8.03 | 0.000 |
| New York | 26.2963 | 3.2417 | 8.11 | 0.000 |
| Ohio | 26.3468 | 3.2608 | 8.08 | 0.000 |
| Oklahoma | 25.8892 | 3.2644 | 7.93 | 0.000 |
| Oregon | 25.4513 | 3.2582 | 7.81 | 0.000 |
| Pennsylvania | 26.4789 | 3.2573 | 8.13 | 0.000 |
| Rhode Island | 25.1521 | 3.2683 | 8.00 | 0.000 |
| South Carolina | 26.4216 | 3.2433 | 8.15 | 0.000 |
| South Dakota | 25.4075 | 3.2665 | 7.78 | 0.000 |
| Tennessee | 26.0621 | 3.2686 | 8.02 | 0.000 |
| Texas | 26.8796 | 3.2213 | 8.34 | 0.000 |
| Utah | 25.7632 | 3.2536 | 7.92 | 0.000 |
| Virginia | 26.3402 | 3.2635 | 8.07 | 0.000 |
| Vermont | 24.3453 | 3.2683 | 7.45 | 0.000 |
| Washington | 26.0738 | 3.2581 | 8.00 | 0.000 |
| Wisconsin | 25.9419 | 3.2617 | 7.95 | 0.000 |
| West Virginia | 25.2368 | 3.2718 | 7.71 | 0.000 |
| Wyoming | 23.9943 | 3.2677 | 7.34 | 0.000 |
| LogDays<sub>t</sub> | -10.4430 | 1.5361 | -6.80 | 0.000 |
| LogDays<sub>t</sub>*LogDays<sub>t</sub> | 1.1916 | 0.1799 | 6.30 | 0.000 |
| LogDailyDead<sub>it-7</sub> | 0.8697 | 0.0246 | 35.32 | 0.000 |
| LogMaxTemp<sub>it-7</sub> | 0.2988 | 0.0506 | 5.91 | 0.000 |
| LogMaxTemp<sub>it-14</sub> | 0.3011 | 0.0545 | 5.52 | 0.000 |
| R<sup>2</sup> | 0.9667 |  |  |  |
| Root MSE | 9.3198 |  |  |  |
| Observations | 4567 |  |  |  |

**Table A2.**
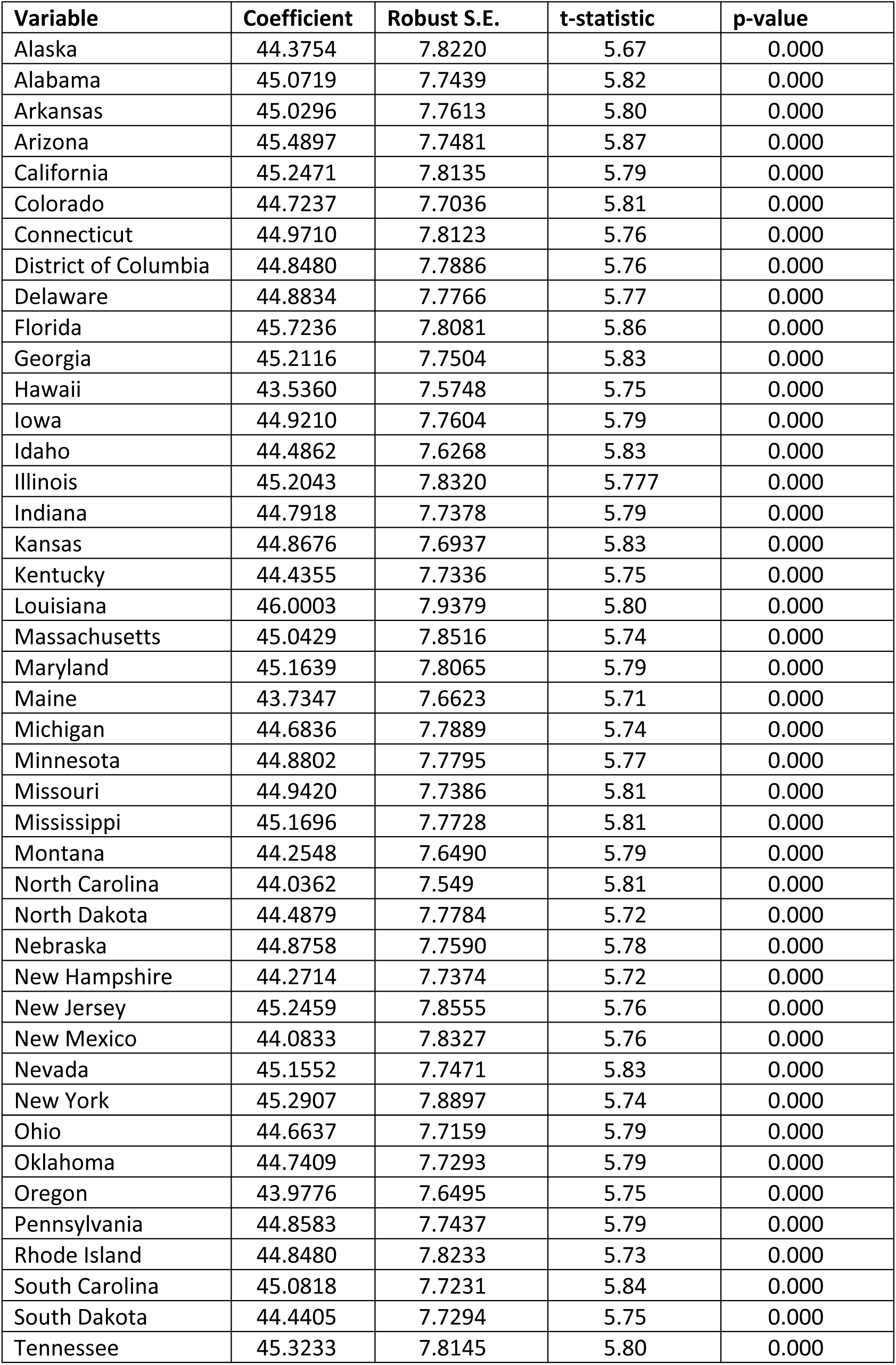
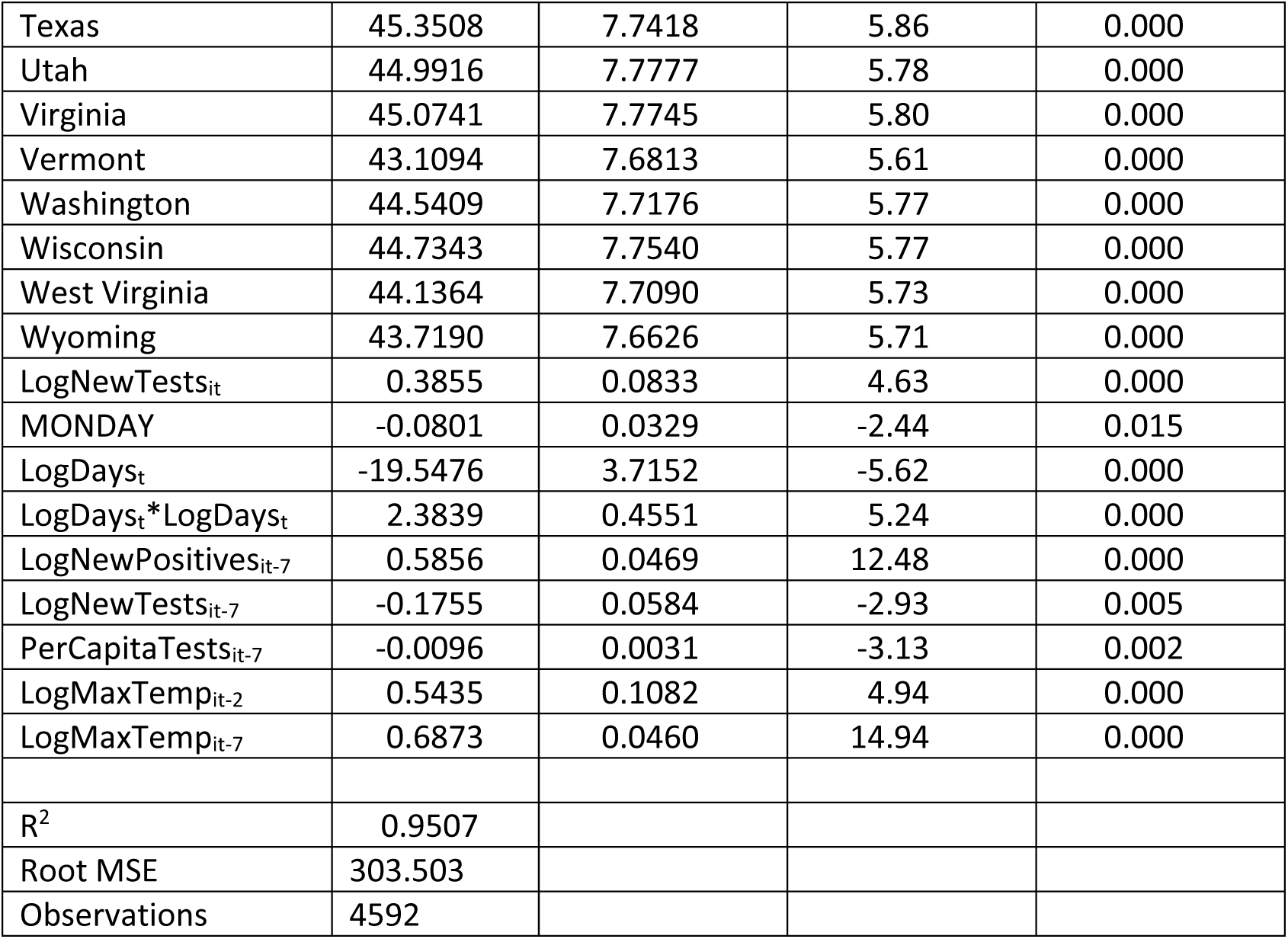
Model predicting NewPositives_it_.

| **Variable** | **Coefficient** | **Robust S.E.** | **t-statistic** | **p-value** |
| --- | --- | --- | --- | --- |
| Alaska | 44.3754 | 7.8220 | 5.67 | 0.000 |
| Alabama | 45.0719 | 7.7439 | 5.82 | 0.000 |
| Arkansas | 45.0296 | 7.7613 | 5.80 | 0.000 |
| Arizona | 45.4897 | 7.7481 | 5.87 | 0.000 |
| California | 45.2471 | 7.8135 | 5.79 | 0.000 |
| Colorado | 44.7237 | 7.7036 | 5.81 | 0.000 |
| Connecticut | 44.9710 | 7.8123 | 5.76 | 0.000 |
| District of Columbia | 44.8480 | 7.7886 | 5.76 | 0.000 |
| Delaware | 44.8834 | 7.7766 | 5.77 | 0.000 |
| Florida | 45.7236 | 7.8081 | 5.86 | 0.000 |
| Georgia | 45.2116 | 7.7504 | 5.83 | 0.000 |
| Hawaii | 43.5360 | 7.5748 | 5.75 | 0.000 |
| Iowa | 44.9210 | 7.7604 | 5.79 | 0.000 |
| Idaho | 44.4862 | 7.6268 | 5.83 | 0.000 |
| Illinois | 45.2043 | 7.8320 | 5.777 | 0.000 |
| Indiana | 44.7918 | 7.7378 | 5.79 | 0.000 |
| Kansas | 44.8676 | 7.6937 | 5.83 | 0.000 |
| Kentucky | 44.4355 | 7.7336 | 5.75 | 0.000 |
| Louisiana | 46.0003 | 7.9379 | 5.80 | 0.000 |
| Massachusetts | 45.0429 | 7.8516 | 5.74 | 0.000 |
| Maryland | 45.1639 | 7.8065 | 5.79 | 0.000 |
| Maine | 43.7347 | 7.6623 | 5.71 | 0.000 |
| Michigan | 44.6836 | 7.7889 | 5.74 | 0.000 |
| Minnesota | 44.8802 | 7.7795 | 5.77 | 0.000 |
| Missouri | 44.9420 | 7.7386 | 5.81 | 0.000 |
| Mississippi | 45.1696 | 7.7728 | 5.81 | 0.000 |
| Montana | 44.2548 | 7.6490 | 5.79 | 0.000 |
| North Carolina | 44.0362 | 7.549 | 5.81 | 0.000 |
| North Dakota | 44.4879 | 7.7784 | 5.72 | 0.000 |
| Nebraska | 44.8758 | 7.7590 | 5.78 | 0.000 |
| New Hampshire | 44.2714 | 7.7374 | 5.72 | 0.000 |
| New Jersey | 45.2459 | 7.8555 | 5.76 | 0.000 |
| New Mexico | 44.0833 | 7.8327 | 5.76 | 0.000 |
| Nevada | 45.1552 | 7.7471 | 5.83 | 0.000 |
| New York | 45.2907 | 7.8897 | 5.74 | 0.000 |
| Ohio | 44.6637 | 7.7159 | 5.79 | 0.000 |
| Oklahoma | 44.7409 | 7.7293 | 5.79 | 0.000 |
| Oregon | 43.9776 | 7.6495 | 5.75 | 0.000 |
| Pennsylvania | 44.8583 | 7.7437 | 5.79 | 0.000 |
| Rhode Island | 44.8480 | 7.8233 | 5.73 | 0.000 |
| South Carolina | 45.0818 | 7.7231 | 5.84 | 0.000 |
| South Dakota | 44.4405 | 7.7294 | 5.75 | 0.000 |
| Tennessee | 45.3233 | 7.8145 | 5.80 | 0.000 |
| Texas | 45.3508 | 7.7418 | 5.86 | 0.000 |
| Utah | 44.9916 | 7.7777 | 5.78 | 0.000 |
| Virginia | 45.0741 | 7.7745 | 5.80 | 0.000 |
| Vermont | 43.1094 | 7.6813 | 5.61 | 0.000 |
| Washington | 44.5409 | 7.7176 | 5.77 | 0.000 |
| Wisconsin | 44.7343 | 7.7540 | 5.77 | 0.000 |
| West Virginia | 44.1364 | 7.7090 | 5.73 | 0.000 |
| Wyoming | 43.7190 | 7.6626 | 5.71 | 0.000 |
| LogNewTests<sub>it</sub> | 0.3855 | 0.0833 | 4.63 | 0.000 |
| MONDAY | -0.0801 | 0.0329 | -2.44 | 0.015 |
| LogDays<sub>t</sub> | -19.5476 | 3.7152 | -5.62 | 0.000 |
| LogDays<sub>t</sub>*LogDays<sub>t</sub> | 2.3839 | 0.4551 | 5.24 | 0.000 |
| LogNewPositives<sub>it-7</sub> | 0.5856 | 0.0469 | 12.48 | 0.000 |
| LogNewTests<sub>it-7</sub> | -0.1755 | 0.0584 | -2.93 | 0.005 |
| PerCapitaTests<sub>it-7</sub> | -0.0096 | 0.0031 | -3.13 | 0.002 |
| LogMaxTemp<sub>it-2</sub> | 0.5435 | 0.1082 | 4.94 | 0.000 |
| LogMaxTemp<sub>it-7</sub> | 0.6873 | 0.0460 | 14.94 | 0.000 |
| R<sup>2</sup> | 0.9507 |  |  |  |
| Root MSE | 303.503 |  |  |  |
| Observations | 4592 |  |  |  |

**Table A3.** Model predicting StateBase_i_ taken from DailyDead_it_ model.

| Variable | Coefficient | Robust S.E. | t-statistic | p-value |
| --- | --- | --- | --- | --- |
| Constant | -2.6082 | 0.2842 | -9.18 | 0.000 |
| LogPopulation | 0.3652 | 0.0430 | 8.48 | 0.000 |
| %Black | 0.0197 | 0.0030 | 6.52 | 0.000 |
| %Hispanic | 0.0172 | 0.0049 | 3.54 | 0.001 |
| %Age 80+ | 0.1163 | 0.0627 | 1.86 | 0.070 |
| R<sup>2</sup> | 0.7938 |  |  |  |
| Root MSE | 0.2845 |  |  |  |
| Observations | 51 |  |  |  |

**Table A4.** Alternative specifications for DailyDead_it_ model. StateIndicator_i_ for models provided in Table A6. Robust standard errors clustered at the state level.

| Variable | Eq. 1<br>Base<br>Model | Eq. 3<br>Linear<br>MaxTemp | Eq. 4<br>Ratio<br>Scaler | Eq. 5<br>Ratio Scaler<br>Linear<br>MaxTemp | Eq. 6<br>14<sup>th</sup> Lag<br>DailyDead | Eq. 7<br>Rolling<br>Weekly | Eq. 12<br>Add<br>UV |
| --- | --- | --- | --- | --- | --- | --- | --- |
| LogDays<sub>t</sub> | -10.4430 | -10.5842 | -10.2210 | -10.1283 | -30.2125 | -13.9640 | -8.7000 |
|  | (1.5361) | (1.5407) | (1.5499) | (1.5545) | (2.3673) | (1.2216) | (1.3241) |
| LogDays<sub>t</sub>*LogDays<sub>t</sub> | 1.1916 | 1.2093 | 1.1627 | 1.1524 | 3.4253 | 1.5895 | 1.0093 |
|  | (0.1799) | (0.1806) | (0.1810) | (0.1815) | (0.2850) | (0.1453) | (0.1565) |
| LogDailyDead<sub>it-7</sub> | 0.8697 | 0.8681 | 0.8738 | 0.8752 |  |  | 0.8585 |
|  | (0.0246) | (0.0248) | (0.0243) | (0.0243) |  |  | (0.0256) |
| LogDailyDead<sub>it-14</sub> |  |  |  |  | 0.8199 |  |  |
|  |  |  |  |  | (0.0478) |  |  |
| LogWeeklyDead<sub>it-7</sub> |  |  |  |  |  | 0.9280 |  |
|  |  |  |  |  |  | (0.0373) |  |
| LogMaxTemp<sub>it-7</sub> | 0.2988 |  | -0.5907 |  | 0.1843 |  | 0.1364 |
|  | (0.0506) |  | (0.1019) |  | (0.1651) |  | (0.1326) |
| LogMaxTemp<sub>it-14</sub> | 0.3011 |  | -0.5790 |  | 0.4681 |  | 0.2487 |
|  | (0.0545) |  | (0.0887) |  | (0.0726) |  | (0.0578) |
| MaxTemp<sub>it-7</sub> |  | 0.1131 |  | -1.3414 |  |  |  |
|  |  | (0.0190) |  | (0.2704) |  |  |  |
| MaxTemp<sub>it-14</sub> |  | 0.1160 |  | -1.3016 |  |  |  |
|  |  | (0.0222) |  | (0.2224) |  |  |  |
| LogMaxTemp<sub>it-14</sub> |  |  |  |  |  | 0.2215 |  |
|  |  |  |  |  |  | (0.0454) |  |
| LogMaxTemp<sub>it-18</sub> |  |  |  |  |  | 0.3292 |  |
|  |  |  |  |  |  | (0.0523) |  |
|  |  |  |  |  |  |  | 0.5213 |
|  |  |  |  |  |  |  | (0.2100) |
|  |  |  |  |  |  |  | 0.2962 |
|  |  |  |  |  |  |  | (0.1318) |
| R<sup>2</sup> | 0.9667 | 0.9667 | 0.9666 | 0.9666 | 0.9425 | 0.9903 | 0.9674 |
| Root MSE | 9.3198 | 9.3151 | 9.3322 | 9.3394 | 12.2483 | 37.8452 | 9.2235 |
| Observations | 4567 | 4567 | 4567 | 4567 | 4576 | 4552 | 4567 |

**Table A5.** AR(7) DailyDead_it_ models using different Reported/Actual combinations. U.S. states: April 16-July 15. June 25 NJ death count (1877) set to missing in reported. (Robust standard errors clustered at state level).

|  | **Eq. 18**<br>Reported/Reported | **Eq. 19**<br>Actual/Reported | **Eq. 20**<br>Reported/Actual | **Eq. 21**<br>Actual/Actual |
| --- | --- | --- | --- | --- |
| DailyDead<sub>it-7</sub> | 0.6986 | 0.6681 | 0.7360 | 0.7854 |
|  | (0.0069) | (0.0067) | (0.0064) | (0.0035) |
| Constant | 5.0046 | 6.6079 | 3.4596 | 3.1725 |
|  | (0.4210) | (0.4096) | (0.3862) | (0.2138) |
| R<sup>2</sup> | 0.6893 | 0.6820 | 0.7428 | 0.9148 |
| RMSE | 26.545 | 25.828 | 24.154 | 13.371 |
| Observations | 4639 | 4639 | 4639 | 4639 |

**Table A6.** Parameter estimates including state-level fixed effects for all models.

| **Model 1** |  |  | **Model 2** |  |  | **Model 3** |  |  |
| --- | --- | --- | --- | --- | --- | --- | --- | --- |
| Variable | Coefficient | S.E. | Variable | Coefficient | S.E. | Variable | Coefficient | S.E. |
| State_AK | 24.2468 | 3.2668 | LogPopulation | 110.6656 | 5.0218 | State_AK | 24.5529 | 3.2749 |
| State_AL | 26.2830 | 3.2504 | Good_Dates | -58.9392 | 10.3053 | State_AL | 26.6085 | 3.2602 |
| State_AR | 25.6412 | 3.2496 | _cons | -799.2542 | 40.5126 | State_AR | 25.9674 | 3.2590 |
| State_AZ | 26.6399 | 3.2277 |  |  |  | State_AZ | 26.9905 | 3.2380 |
| State_CA | 26.5480 | 3.2434 | R-square | 0.0965 |  | State_CA | 26.8647 | 3.2529 |
| State_CO | 26.1724 | 3.2608 | Root MSE | 330.1528 |  | State_CO | 26.4937 | 3.2708 |
| State_CT | 26.3270 | 3.2578 | Observations | 4567 |  | State_CT | 26.6431 | 3.2674 |
| State_DC | 26.0314 | 3.2699 |  |  |  | State_DC | 26.3474 | 3.2795 |
| State_DE | 26.0525 | 3.2719 |  |  |  | State_DE | 26.3659 | 3.2813 |
| State_FL | 26.7896 | 3.2194 |  |  |  | State_FL | 27.1275 | 3.2294 |
| State_GA | 26.4536 | 3.2545 |  |  |  | State_GA | 26.7781 | 3.2646 |
| State_HI | 23.8933 | 3.2676 |  |  |  | State_HI | 24.2219 | 3.2771 |
| State_IA | 26.1666 | 3.2669 |  |  |  | State_IA | 26.4856 | 3.2764 |
| State_ID | 25.3286 | 3.2630 |  |  |  | State_ID | 25.6418 | 3.2723 |
| State_IL | 26.5186 | 3.2569 |  |  |  | State_IL | 26.8400 | 3.2668 |
| State_IN | 26.3159 | 3.2619 |  |  |  | State_IN | 26.6345 | 3.2716 |
| State_KS | 25.6968 | 3.2630 |  |  |  | State_KS | 26.0204 | 3.2729 |
| State_KY | 25.8271 | 3.2613 |  |  |  | State_KY | 26.1429 | 3.2709 |
| State_LA | 26.3165 | 3.2548 |  |  |  | State_LA | 26.6510 | 3.2655 |
| State_MA | 26.4245 | 3.2557 |  |  |  | State_MA | 26.7435 | 3.2653 |
| State_MD | 26.4151 | 3.2614 |  |  |  | State_MD | 26.7333 | 3.2713 |
| State_ME | 25.0056 | 3.2684 |  |  |  | State_ME | 25.3192 | 3.2773 |
| State_MI | 26.2274 | 3.2515 |  |  |  | State_MI | 26.5469 | 3.2613 |
| State_MN | 26.3019 | 3.2628 |  |  |  | State_MN | 26.6185 | 3.2723 |
| State_MO | 25.9973 | 3.2607 |  |  |  | State_MO | 26.3193 | 3.2706 |
| State_MS | 26.2978 | 3.2530 |  |  |  | State_MS | 26.6264 | 3.2628 |
| State_MT | 24.7679 | 3.2679 |  |  |  | State_MT | 25.0807 | 3.2770 |
| State_NC | 26.3227 | 3.2556 |  |  |  | State_NC | 26.6467 | 3.2654 |
| State_ND | 25.2256 | 3.2686 |  |  |  | State_ND | 25.5440 | 3.2777 |
| State_NE | 25.7297 | 3.2615 |  |  |  | State_NE | 26.0525 | 3.2711 |
| State_NH | 25.8414 | 3.2635 |  |  |  | State_NH | 26.1593 | 3.2730 |
| State_NJ | 26.3998 | 3.2477 |  |  |  | State_NJ | 26.7177 | 3.2579 |
| State_NM | 26.1024 | 3.2650 |  |  |  | State_NM | 26.4317 | 3.2749 |
| State_NV | 26.1382 | 3.2566 |  |  |  | State_NV | 26.4738 | 3.2667 |
| State_NY | 26.2963 | 3.2417 |  |  |  | State_NY | 26.6136 | 3.2519 |
| State_OH | 26.3468 | 3.2608 |  |  |  | State_OH | 26.6642 | 3.2703 |
| State_OK | 25.8892 | 3.2644 |  |  |  | State_OK | 26.2148 | 3.2743 |
| State_OR | 25.4513 | 3.2582 |  |  |  | State_OR | 25.7649 | 3.2675 |
| State_PA | 26.4789 | 3.2573 |  |  |  | State_PA | 26.7961 | 3.2673 |
| State_RI | 26.1521 | 3.2683 |  |  |  | State_RI | 26.4658 | 3.2776 |
| State_SC | 26.4216 | 3.2433 |  |  |  | State_SC | 26.7494 | 3.2530 |
| State_SD | 25.4075 | 3.2665 |  |  |  | State_SD | 25.7246 | 3.2758 |
| State_TN | 26.0621 | 3.2486 |  |  |  | State_TN | 26.3870 | 3.2582 |
| State_TX | 26.8796 | 3.2213 |  |  |  | State_TX | 27.2139 | 3.2313 |
| State_UT | 25.7632 | 3.2536 |  |  |  | State_UT | 26.0825 | 3.2629 |
| State_VA | 26.3402 | 3.2635 |  |  |  | State_VA | 26.6573 | 3.2733 |
| State_VT | 24.3453 | 3.2683 |  |  |  | State_VT | 24.6667 | 3.2772 |
| State_WA | 26.0738 | 3.2581 |  |  |  | State_WA | 26.3857 | 3.2676 |
| State_WI | 25.9419 | 3.2617 |  |  |  | State_WI | 26.2584 | 3.2708 |
| State_WV | 25.2368 | 3.2718 |  |  |  | State_WV | 25.5517 | 3.2811 |
| State_WY | 23.9943 | 3.2677 |  |  |  | State_WY | 24.3020 | 3.2761 |
| t0 | -10.4430 | 1.5361 |  |  |  | t0 | -10.5842 | 1.5407 |
| t2 | 1.1916 | 0.1799 |  |  |  | t2 | 1.2093 | 0.1806 |
| b1 | 0.8697 | 0.0246 |  |  |  | b1 | 0.8681 | 0.0248 |
| w1 | 0.2988 | 0.0506 |  |  |  | k1 | 0.1131 | 0.0190 |
| w2 | 0.3011 | 0.0545 |  |  |  | k2 | 0.1160 | 0.0222 |
| R-square | 0.9667 |  |  |  |  | R-square | 0.9667 |  |
| Root MSE | 9.3198 |  |  |  |  | Root MSE | 9.3152 |  |
| Observations | 4567 |  |  |  |  | Observations | 4567 |  |

**Model 4**
| Variable | Coefficient | S.E. |
| --- | --- | --- |
| State_AK | 20.1337 | 3.3174 |
| State_AL | 22.1346 | 3.2978 |
| State_AR | 21.4935 | 3.2976 |
| State_AZ | 22.4619 | 3.2750 |
| State_CA | 22.4087 | 3.2909 |
| State_CO | 22.0283 | 3.3069 |
| State_CT | 22.1941 | 3.3051 |
| State_DC | 21.8982 | 3.3169 |
| State_DE | 21.9243 | 3.3194 |
| State_FL | 22.6230 | 3.2668 |
| State_GA | 22.3049 | 3.3007 |
| State_HI | 19.7449 | 3.3147 |
| State_IA | 22.0278 | 3.3149 |
| State_ID | 21.2024 | 3.3106 |
| State_IL | 22.3739 | 3.3042 |
| State_IN | 22.1782 | 3.3090 |
| State_KS | 21.5547 | 3.3097 |
| State_KY | 21.6948 | 3.3089 |
| State_LA | 22.1540 | 3.2993 |
| State_MA | 22.2860 | 3.3037 |
| State_MD | 22.2768 | 3.3081 |
| State_ME | 20.8797 | 3.3174 |
| State_MI | 22.0856 | 3.2989 |
| State_MN | 22.1667 | 3.3107 |
| State_MO | 21.8541 | 3.3075 |
| State_MS | 22.1459 | 3.3003 |
| State_MT | 20.6421 | 3.3164 |
| State_NC | 22.1764 | 3.3026 |
| State_ND | 21.0901 | 3.3174 |
| State_NE | 21.5877 | 3.3092 |
| State_NH | 21.7067 | 3.3115 |
| State_NJ | 22.2613 | 3.2935 |
| State_NM | 21.9509 | 3.3120 |
| State_NV | 21.9790 | 3.3031 |
| State_NY | 22.1590 | 3.2876 |
| State_OH | 22.2103 | 3.3087 |
| State_OK | 21.7425 | 3.3109 |
| State_OR | 21.3224 | 3.3057 |
| State_PA | 22.3421 | 3.3038 |
| State_RI | 22.0239 | 3.3165 |
| State_SC | 22.2712 | 3.2909 |
| State_SD | 21.2743 | 3.3149 |
| State_TN | 21.9162 | 3.2961 |
| State_TX | 22.7186 | 3.2689 |
| State_UT | 21.6275 | 3.3019 |
| State_VA | 22.2043 | 3.3103 |
| State_VT | 20.2029 | 3.3185 |
| State_WA | 21.9464 | 3.3051 |
| State_WI | 21.8095 | 3.3107 |
| State_WV | 21.1075 | 3.3194 |
| State_WY | 19.8756 | 3.3181 |
| t0 | -10.2110 | 1.5499 |
| t2 | 1.1627 | 0.1810 |
| b1 | 0.8738 | 0.0243 |
| a1 | -0.5907 | 0.1019 |
| a2 | -0.5790 | 0.0887 |
| R-square | 0.9666 |  |
| Root MSE | 9.3322 |  |
| Observations | 4567 |  |

**Model 5**
| Variable | Coefficient | S.E. |
| --- | --- | --- |
| State_AK | 20.2699 | 3.3231 |
| State_AL | 22.2581 | 3.3039 |
| State_AR | 21.6173 | 3.3036 |
| State_AZ | 22.5749 | 3.2812 |
| State_CA | 22.5354 | 3.2970 |
| State_CO | 22.1544 | 3.3129 |
| State_CT | 22.3229 | 3.3110 |
| State_DC | 22.0268 | 3.3228 |
| State_DE | 22.0545 | 3.3253 |
| State_FL | 22.7402 | 3.2728 |
| State_GA | 22.4284 | 3.3067 |
| State_HI | 19.8683 | 3.3209 |
| State_IA | 22.1551 | 3.3209 |
| State_ID | 21.3334 | 3.3165 |
| State_IL | 22.4993 | 3.3102 |
| State_IN | 22.3055 | 3.3149 |
| State_KS | 21.6802 | 3.3157 |
| State_KY | 21.8238 | 3.3147 |
| State_LA | 22.2725 | 3.3055 |
| State_MA | 22.4136 | 3.3096 |
| State_MD | 22.4036 | 3.3140 |
| State_ME | 21.0117 | 3.3232 |
| State_MI | 22.2125 | 3.3050 |
| State_MN | 22.2947 | 3.3166 |
| State_MO | 21.9793 | 3.3134 |
| State_MS | 22.2682 | 3.3064 |
| State_MT | 20.7733 | 3.3223 |
| State_NC | 22.3006 | 3.3086 |
| State_ND | 21.2190 | 3.3233 |
| State_NE | 21.7133 | 3.3151 |
| State_NH | 21.8348 | 3.3174 |
| State_NJ | 22.3879 | 3.2995 |
| State_NM | 22.0732 | 3.3181 |
| State_NV | 22.0985 | 3.3093 |
| State_NY | 22.2860 | 3.2936 |
| State_OH | 22.3383 | 3.3147 |
| State_OK | 21.8670 | 3.3169 |
| State_OR | 21.4524 | 3.3116 |
| State_PA | 22.4694 | 3.3097 |
| State_RI | 22.1545 | 3.3224 |
| State_SC | 22.3941 | 3.2969 |
| State_SD | 21.4031 | 3.3208 |
| State_TN | 22.0405 | 3.3021 |
| State_TX | 22.8377 | 3.2749 |
| State_UT | 21.7553 | 3.3078 |
| State_VA | 22.3319 | 3.3163 |
| State_VT | 20.3324 | 3.3245 |
| State_WA | 22.0770 | 3.3110 |
| State_WI | 21.9391 | 3.3166 |
| State_WV | 21.2375 | 3.3253 |
| State_WY | 20.0103 | 3.3239 |
| t0 | -10.1283 | 1.5545 |
| t2 | 1.1524 | 0.1815 |
| b1 | 0.8752 | 0.0243 |
| j1 | -1.3414 | 0.2704 |
| j2 | -1.3016 | 0.2224 |
| R-square | 0.9666 |  |
| Root MSE | 9.3394 |  |
| Observations | 4567 |  |

**Model 6**
| Variable | Coefficient | S.E. |
| --- | --- | --- |
| State_AK | 68.0983 | 4.8708 |
| State_AL | 70.0627 | 4.8869 |
| State_AR | 69.3268 | 4.8655 |
| State_AZ | 70.5461 | 4.8661 |
| State_CA | 70.4218 | 4.9037 |
| State_CO | 69.8248 | 4.9467 |
| State_CT | 70.1264 | 4.9560 |
| State_DC | 69.8024 | 4.9293 |
| State_DE | 69.8608 | 4.9324 |
| State_FL | 70.6838 | 4.8716 |
| State_GA | 70.1762 | 4.9259 |
| State_HI | 67.5865 | 4.9040 |
| State_IA | 69.8836 | 4.9124 |
| State_ID | 68.7233 | 4.9201 |
| State_IL | 70.3744 | 4.9345 |
| State_IN | 70.0431 | 4.9376 |
| State_KS | 69.1838 | 4.9171 |
| State_KY | 69.5242 | 4.9171 |
| State_LA | 69.9121 | 4.9466 |
| State_MA | 70.3122 | 4.9548 |
| State_MD | 70.2614 | 4.9348 |
| State_ME | 68.7766 | 4.9195 |
| State_MI | 69.8819 | 4.9552 |
| State_MN | 70.0764 | 4.9105 |
| State_MO | 69.7402 | 4.9144 |
| State_MS | 70.1096 | 4.9029 |
| State_MT | 68.5028 | 4.8888 |
| State_NC | 70.0974 | 4.9015 |
| State_ND | 68.8297 | 4.9008 |
| State_NE | 69.3169 | 4.8851 |
| State_NH | 69.4363 | 4.8960 |
| State_NJ | 70.1194 | 4.9633 |
| State_NM | 69.8534 | 4.9026 |
| State_NV | 69.7741 | 4.9211 |
| State_NY | 69.9672 | 4.9672 |
| State_OH | 70.1541 | 4.9305 |
| State_OK | 69.5545 | 4.9309 |
| State_OR | 69.2116 | 4.9088 |
| State_PA | 70.3375 | 4.9543 |
| State_RI | 69.9466 | 4.9329 |
| State_SC | 70.1774 | 4.8797 |
| State_SD | 69.0783 | 4.8834 |
| State_TN | 69.6894 | 4.8964 |
| State_TX | 70.8674 | 4.8732 |
| State_UT | 69.4429 | 4.8863 |
| State_VA | 70.1628 | 4.9326 |
| State_VT | 67.8890 | 4.9203 |
| State_WA | 69.4145 | 4.9331 |
| State_WI | 69.4335 | 4.9177 |
| State_WV | 68.9390 | 4.9132 |
| State_WY | 67.3690 | 4.8829 |
| t0 | -30.2125 | 2.3673 |
| t2 | 3.4253 | 0.2850 |
| b1 | 0.8199 | 0.0478 |
| d1 | 0.1843 | 0.1651 |
| d2 | 0.4681 | 0.0726 |
| R-square | 0.9425 |  |
| Root MSE | 12.2483 |  |
| Observations | 4567 |  |

**Model 7**
| Variable | Coefficient | S.E. |
| --- | --- | --- |
| State_AK | 32.9102 | 2.6235 |
| State_AL | 34.0669 | 2.7254 |
| State_AR | 33.8438 | 2.6948 |
| State_AZ | 34.2783 | 2.7475 |
| State_CA | 34.1784 | 2.7625 |
| State_CO | 33.8629 | 2.7479 |
| State_CT | 33.9715 | 2.7669 |
| State_DC | 33.9732 | 2.7261 |
| State_DE | 33.9674 | 2.7253 |
| State_FL | 34.3485 | 2.7639 |
| State_GA | 34.1048 | 2.7569 |
| State_HI | 32.9555 | 2.6727 |
| State_IA | 34.0259 | 2.7253 |
| State_ID | 33.3378 | 2.6796 |
| State_IL | 34.1271 | 2.7718 |
| State_IN | 34.0090 | 2.7508 |
| State_KS | 33.7194 | 2.7060 |
| State_KY | 33.8415 | 2.7138 |
| State_LA | 33.9520 | 2.7729 |
| State_MA | 34.0373 | 2.7750 |
| State_MD | 34.0977 | 2.7607 |
| State_ME | 33.4256 | 2.6726 |
| State_MI | 33.8339 | 2.7745 |
| State_MN | 34.0356 | 2.7370 |
| State_MO | 33.9426 | 2.7337 |
| State_MS | 34.0962 | 2.7307 |
| State_MT | 33.4258 | 2.6469 |
| State_NC | 34.0766 | 2.7367 |
| State_ND | 33.6488 | 2.6754 |
| State_NE | 33.7940 | 2.6973 |
| State_NH | 33.8766 | 2.7025 |
| State_NJ | 33.9433 | 2.7998 |
| State_NM | 34.0438 | 2.7222 |
| State_NV | 33.9905 | 2.7232 |
| State_NY | 33.8453 | 2.8111 |
| State_OH | 34.0392 | 2.7488 |
| State_OK | 33.7927 | 2.7157 |
| State_OR | 33.7039 | 2.6892 |
| State_PA | 34.0933 | 2.7813 |
| State_RI | 33.9590 | 2.7275 |
| State_SC | 34.1241 | 2.7177 |
| State_SD | 33.6997 | 2.6770 |
| State_TN | 33.9789 | 2.7113 |
| State_TX | 34.4291 | 2.7524 |
| State_UT | 33.9061 | 2.6837 |
| State_VA | 34.0636 | 2.7508 |
| State_VT | 33.0951 | 2.6638 |
| State_WA | 33.7650 | 2.7259 |
| State_WI | 33.7719 | 2.7117 |
| State_WV | 33.6192 | 2.6880 |
| State_WY | 33.1590 | 2.6433 |
| t0 | -13.9640 | 1.2216 |
| t2 | 1.5895 | 0.1453 |
| b1 | 0.9280 | 0.0373 |
| h1 | 0.2215 | 0.0454 |
| h2 | 0.3292 | 0.0523 |
| R-square | 0.9903 |  |
| Root MSE | 37.8452 |  |
| Observations | 4552 |  |

**Model 8**
| Variable | Coefficient | S.E. |
| --- | --- | --- |
| State_AK | 35.3125 | 4.9526 |
| State_AL | 37.5093 | 5.0265 |
| State_AR | 39.9268 | 6.7158 |
| State_AZ | 37.7874 | 4.9858 |
| State_CA | 37.5128 | 4.9174 |
| State_CO | 37.2238 | 4.9732 |
| State_CT | 37.3223 | 4.9562 |
| State_DC | 37.1079 | 5.0228 |
| State_DE | 36.9748 | 4.9447 |
| State_FL | 38.0022 | 5.0012 |
| State_GA | 37.7061 | 5.0454 |
| State_HI | 34.8410 | 4.9367 |
| State_IA | 40.4395 | 6.6952 |
| State_ID | 36.2677 | 4.9392 |
| State_IL | 37.5012 | 4.9429 |
| State_IN | 37.3513 | 4.9613 |
| State_KS | 36.7697 | 4.9970 |
| State_KY | 36.8543 | 4.9687 |
| State_LA | 37.2923 | 4.9540 |
| State_MA | 37.4709 | 4.9703 |
| State_MD | 37.5591 | 5.0236 |
| State_ME | 36.1441 | 5.0254 |
| State_MI | 37.2267 | 4.9576 |
| State_MN | 37.3694 | 4.9796 |
| State_MO | 37.2742 | 5.0676 |
| State_MS | 37.4815 | 5.0134 |
| State_MT | 35.7994 | 4.9576 |
| State_NC | 37.4502 | 5.0010 |
| State_ND | 39.5098 | 6.7049 |
| State_NE | 40.0185 | 6.7035 |
| State_NH | 36.8973 | 4.9704 |
| State_NJ | 37.3833 | 4.9410 |
| State_NM | 37.0723 | 4.9427 |
| State_NV | 37.2553 | 5.0116 |
| State_NY | 37.3281 | 4.9554 |
| State_OH | 37.3592 | 4.9516 |
| State_OK | 39.8591 | 6.5531 |
| State_OR | 36.3622 | 4.9248 |
| State_PA | 37.7060 | 5.0665 |
| State_RI | 37.2153 | 4.9853 |
| State_SC | 37.6689 | 5.0245 |
| State_SD | 39.7644 | 6.7388 |
| State_TN | 37.2144 | 4.9961 |
| State_TX | 38.0718 | 4.9929 |
| State_UT | 39.8857 | 6.6641 |
| State_VA | 37.4447 | 5.0106 |
| State_VT | 35.2688 | 4.9384 |
| State_WA | 36.9972 | 4.9326 |
| State_WI | 36.9340 | 4.9507 |
| State_WV | 36.1957 | 4.9344 |
| State_WY | 38.3688 | 6.7926 |
| t0 | -15.8165 | 2.4326 |
| t2 | 1.6788 | 0.2216 |
| s1 | 0.8278 | 0.5701 |
| s2 | -0.0520 | 0.0253 |
| b1 | 0.8318 | 0.0247 |
| w1 | 0.2903 | 0.0482 |
| w2 | 0.2849 | 0.0495 |
| R-square | 0.9677 |  |
| Root MSE | 9.1764 |  |
| Observations | 4567 |  |

**Model 9**
| Variable | Coefficient | S.E. |
| --- | --- | --- |
| State_AK | 1.8320 | 0.3592 |
| State_AL | 3.6712 | 0.4368 |
| State_AR | 2.3075 | 0.1980 |
| State_AZ | 4.0871 | 0.5048 |
| State_CA | 3.9824 | 0.5123 |
| State_CO | 3.6027 | 0.4206 |
| State_CT | 3.7619 | 0.4606 |
| State_DC | 3.5902 | 0.3880 |
| State_DE | 3.6529 | 0.4065 |
| State_FL | 4.1604 | 0.5430 |
| State_GA | 3.7659 | 0.4377 |
| State_HI | 1.5444 | 0.3470 |
| State_IA | 2.7473 | 0.1963 |
| State_ID | 2.9995 | 0.3601 |
| State_IL | 3.9148 | 0.5004 |
| State_IN | 3.7293 | 0.4461 |
| State_KS | 3.2786 | 0.3736 |
| State_KY | 3.3538 | 0.4161 |
| State_LA | 3.7792 | 0.4412 |
| State_MA | 3.7945 | 0.4852 |
| State_MD | 3.8027 | 0.4474 |
| State_ME | 2.6020 | 0.3471 |
| State_MI | 3.6610 | 0.4664 |
| State_MN | 3.7560 | 0.4508 |
| State_MO | 3.3604 | 0.4222 |
| State_MS | 3.7118 | 0.4328 |
| State_MT | 2.3461 | 0.3493 |
| State_NC | 3.7650 | 0.4384 |
| State_ND | 1.9761 | 0.1599 |
| State_NE | 2.3871 | 0.1875 |
| State_NH | 3.3717 | 0.4212 |
| State_NJ | 3.7457 | 0.5071 |
| State_NM | 3.6508 | 0.4188 |
| State_NV | 3.6601 | 0.3946 |
| State_NY | 3.5950 | 0.5251 |
| State_OH | 3.7773 | 0.4583 |
| State_OK | 2.5978 | 0.1798 |
| State_OR | 3.1231 | 0.3872 |
| State_PA | 3.7882 | 0.4607 |
| State_RI | 3.6310 | 0.4157 |
| State_SC | 3.8521 | 0.4282 |
| State_SD | 2.0854 | 0.1686 |
| State_TN | 3.5423 | 0.4293 |
| State_TX | 4.2796 | 0.5281 |
| State_UT | 2.5229 | 0.1775 |
| State_VA | 3.7735 | 0.4327 |
| State_VT | 2.1092 | 0.3380 |
| State_WA | 3.6787 | 0.4133 |
| State_WI | 3.5068 | 0.4234 |
| State_WV | 2.8448 | 0.3730 |
| State_WY | 0.8432 | 0.1415 |
| s1 | -0.2059 | 0.0909 |
| s2 | 0.0703 | 0.0183 |
| b1 | 0.9662 | 0.0441 |
| w1 | 0.2678 | 0.0544 |
| w2 | 0.2130 | 0.0771 |
| R-square | 0.9622 |  |
| Root MSE | 9.9251 |  |
| Observations | 4567 |  |

**Model 10**
| Variable | Coefficient | S.E. |
| --- | --- | --- |
| State_AK | 26.8779 | 3.2441 |
| State_AL | 28.9128 | 3.2286 |
| State_AR | 28.2708 | 3.2280 |
| State_AZ | 29.2751 | 3.2005 |
| State_CA | 29.1775 | 3.2217 |
| State_CO | 28.8056 | 3.2357 |
| State_CT | 28.9574 | 3.2353 |
| State_DC | 28.6617 | 3.2477 |
| State_DE | 28.6829 | 3.2497 |
| State_FL | 29.4184 | 3.1982 |
| State_GA | 29.0838 | 3.2322 |
| State_HI | 26.5230 | 3.2463 |
| State_IA | 28.7965 | 3.2449 |
| State_ID | 27.9617 | 3.2382 |
| State_IL | 29.1492 | 3.2343 |
| State_IN | 28.9459 | 3.2399 |
| State_KS | 28.3278 | 3.2402 |
| State_KY | 28.4573 | 3.2392 |
| State_LA | 28.9455 | 3.2337 |
| State_MA | 29.0544 | 3.2337 |
| State_MD | 29.0454 | 3.2391 |
| State_ME | 27.6359 | 3.2462 |
| State_MI | 28.8579 | 3.2290 |
| State_MN | 28.9328 | 3.2397 |
| State_MO | 28.6276 | 3.2382 |
| State_MS | 28.9273 | 3.2314 |
| State_MT | 27.4010 | 3.2435 |
| State_NC | 28.9526 | 3.2336 |
| State_ND | 27.8562 | 3.2459 |
| State_NE | 28.3603 | 3.2390 |
| State_NH | 28.4725 | 3.2400 |
| State_NJ | 29.0300 | 3.2255 |
| State_NM | 28.7391 | 3.2366 |
| State_NV | 28.7745 | 3.2286 |
| State_NY | 28.9253 | 3.2207 |
| State_OH | 28.9768 | 3.2388 |
| State_OK | 28.5192 | 3.2425 |
| State_OR | 28.0822 | 3.2355 |
| State_PA | 29.1095 | 3.2347 |
| State_RI | 28.7823 | 3.2463 |
| State_SC | 29.0509 | 3.2219 |
| State_SD | 28.0382 | 3.2439 |
| State_TN | 28.6923 | 3.2264 |
| State_TX | 29.5088 | 3.1998 |
| State_UT | 28.3967 | 3.2283 |
| State_VA | 28.9706 | 3.2412 |
| State_VT | 26.9756 | 3.2467 |
| State_WA | 28.7046 | 3.2355 |
| State_WI | 28.5722 | 3.2397 |
| State_WV | 27.8668 | 3.2499 |
| State_WY | 26.6265 | 3.2436 |
| t0 | -10.4435 | 1.5277 |
| t2 | 1.1916 | 0.1790 |
| b1 | 0.8698 | 0.0246 |
| w1 | 0.3033 | 0.0680 |
| w2 | 0.3054 | 0.0696 |
| a1 | -0.0079 | 0.0469 |
| a2 | -0.0087 | 0.0460 |
| R-square | 0.9667 |  |
| Root MSE | 9.3217 |  |
| Observations | 4567 |  |

**Model 11**
| Variable | Coefficient | S.E. |
| --- | --- | --- |
| State_AK | 26.9609 | 3.3219 |
| State_AL | 28.9971 | 3.3067 |
| State_AR | 28.3566 | 3.3060 |
| State_AZ | 29.3271 | 3.2730 |
| State_CA | 29.2619 | 3.2998 |
| State_CO | 28.8851 | 3.3155 |
| State_CT | 29.0422 | 3.3141 |
| State_DC | 28.7447 | 3.3246 |
| State_DE | 28.7667 | 3.3274 |
| State_FL | 29.5033 | 3.2764 |
| State_GA | 29.1664 | 3.3097 |
| State_HI | 26.6045 | 3.3212 |
| State_IA | 28.8814 | 3.3228 |
| State_ID | 28.0400 | 3.3156 |
| State_IL | 29.2323 | 3.3130 |
| State_IN | 29.0312 | 3.3180 |
| State_KS | 28.4099 | 3.3172 |
| State_KY | 28.5421 | 3.3170 |
| State_LA | 29.0298 | 3.3095 |
| State_MA | 29.1391 | 3.3117 |
| State_MD | 29.1288 | 3.3173 |
| State_ME | 27.7199 | 3.3232 |
| State_MI | 28.9423 | 3.3073 |
| State_MN | 29.0141 | 3.3178 |
| State_MO | 28.7101 | 3.3157 |
| State_MS | 29.0130 | 3.3094 |
| State_MT | 27.4784 | 3.3209 |
| State_NC | 29.0372 | 3.3116 |
| State_ND | 27.9402 | 3.3240 |
| State_NE | 28.4443 | 3.3174 |
| State_NH | 28.5546 | 3.3183 |
| State_NJ | 29.1130 | 3.3032 |
| State_NM | 28.7988 | 3.3131 |
| State_NV | 28.8261 | 3.2988 |
| State_NY | 29.0122 | 3.2984 |
| State_OH | 29.0620 | 3.3170 |
| State_OK | 28.6046 | 3.3198 |
| State_OR | 28.1656 | 3.3131 |
| State_PA | 29.1915 | 3.3127 |
| State_RI | 28.8678 | 3.3245 |
| State_SC | 29.1369 | 3.3002 |
| State_SD | 28.1215 | 3.3218 |
| State_TN | 28.7762 | 3.3044 |
| State_TX | 29.5947 | 3.2791 |
| State_UT | 28.4689 | 3.3040 |
| State_VA | 29.0544 | 3.3193 |
| State_VT | 27.0602 | 3.3232 |
| State_WA | 28.7882 | 3.3135 |
| State_WI | 28.6559 | 3.3170 |
| State_WV | 27.9526 | 3.3273 |
| State_WY | 26.7093 | 3.3234 |
| t0 | -10.4301 | 1.5426 |
| t2 | 1.1901 | 0.1809 |
| b1 | 0.8696 | 0.0248 |
| w1 | 0.2950 | 0.0568 |
| w2 | 0.2997 | 0.0579 |
| a1 | 0.0571 | 0.1556 |
| a2 | 0.0489 | 0.1600 |
| R-square | 0.9667 |  |
| Root MSE | 9.3210 |  |
| Observations | 4567 |  |

**Model 12**
| Variable | Coefficient | S.E. |
| --- | --- | --- |
| State_AK | 22.6847 | 2.8044 |
| State_AL | 25.1718 | 2.7852 |
| State_AR | 24.5079 | 2.7841 |
| State_AZ | 25.5209 | 2.7633 |
| State_CA | 25.4578 | 2.7795 |
| State_CO | 25.0437 | 2.7958 |
| State_CT | 25.1276 | 2.7915 |
| State_DC | 24.8483 | 2.8024 |
| State_DE | 24.8631 | 2.8045 |
| State_FL | 25.7442 | 2.7570 |
| State_GA | 25.3558 | 2.7893 |
| State_HI | 22.8186 | 2.8026 |
| State_IA | 24.9829 | 2.7995 |
| State_ID | 24.0953 | 2.7966 |
| State_IL | 25.3361 | 2.7902 |
| State_IN | 25.1426 | 2.7954 |
| State_KS | 24.5349 | 2.7971 |
| State_KY | 24.6568 | 2.7946 |
| State_LA | 25.2766 | 2.7917 |
| State_MA | 25.2375 | 2.7903 |
| State_MD | 25.2423 | 2.7944 |
| State_ME | 23.7763 | 2.8028 |
| State_MI | 25.0237 | 2.7851 |
| State_MN | 25.0833 | 2.7955 |
| State_MO | 24.8275 | 2.7937 |
| State_MS | 25.1949 | 2.7877 |
| State_MT | 23.5121 | 2.8019 |
| State_NC | 25.1977 | 2.7895 |
| State_ND | 23.9670 | 2.8017 |
| State_NE | 24.5407 | 2.7947 |
| State_NH | 24.6115 | 2.7965 |
| State_NJ | 25.2245 | 2.7820 |
| State_NM | 25.0030 | 2.7989 |
| State_NV | 24.9894 | 2.7899 |
| State_NY | 25.1382 | 2.7770 |
| State_OH | 25.1625 | 2.7942 |
| State_OK | 24.7573 | 2.7998 |
| State_OR | 24.1740 | 2.7918 |
| State_PA | 25.3055 | 2.7906 |
| State_RI | 24.9457 | 2.8015 |
| State_SC | 25.3066 | 2.7785 |
| State_SD | 24.1938 | 2.7990 |
| State_TN | 24.9281 | 2.7831 |
| State_TX | 25.7912 | 2.7588 |
| State_UT | 24.6060 | 2.7886 |
| State_VA | 25.1686 | 2.7964 |
| State_VT | 23.1031 | 2.8024 |
| State_WA | 24.7820 | 2.7913 |
| State_WI | 24.7357 | 2.7956 |
| State_WV | 24.0587 | 2.8047 |
| State_WY | 22.8521 | 2.8023 |
| t0 | -8.7000 | 1.3241 |
| t2 | 1.0093 | 0.1565 |
| b1 | 0.8585 | 0.0256 |
| w1 | 0.1364 | 0.1326 |
| w2 | 0.2487 | 0.0578 |
| a1 | 0.5213 | 0.2100 |
| a2 | 0.2962 | 0.1318 |
| R-square | 0.9674 |  |
| Root MSE | 9.2236 |  |
| Observations | 4567 |  |

**Model 13**
| Variable | Coefficien | S.E. |
| --- | --- | --- |
| State_AK | 16.9355 | 3.469 |
| State_AL | 19.5465 | 3.481 |
| State_AR | 18.8769 | 3.478 |
| State_AZ | 19.8707 | 3.459 |
| State_CA | 19.8457 | 3.474 |
| State_CO | 19.4279 | 3.485 |
| State_CT | 19.4921 | 3.473 |
| State_DC | 19.2066 | 3.491 |
| State_DE | 19.2292 | 3.492 |
| State_FL | 20.1358 | 3.454 |
| State_GA | 19.7321 | 3.484 |
| State_HI | 17.2231 | 3.502 |
| State_IA | 19.3406 | 3.489 |
| State_ID | 18.4468 | 3.480 |
| State_IL | 19.6902 | 3.477 |
| State_IN | 19.5082 | 3.483 |
| State_KS | 18.8991 | 3.487 |
| State_KY | 19.0255 | 3.482 |
| State_LA | 19.6669 | 3.491 |
| State_MA | 19.6161 | 3.471 |
| State_MD | 19.5957 | 3.483 |
| State_ME | 18.1559 | 3.483 |
| State_MI | 19.3781 | 3.464 |
| State_MN | 19.4252 | 3.482 |
| State_MO | 19.1811 | 3.484 |
| State_MS | 19.5727 | 3.485 |
| State_MT | 17.8518 | 3.486 |
| State_NC | 19.5672 | 3.484 |
| State_ND | 18.3113 | 3.486 |
| State_NE | 18.8928 | 3.485 |
| State_NH | 18.9573 | 3.483 |
| State_NJ | 19.5782 | 3.464 |
| State_NM | 19.3860 | 3.498 |
| State_NV | 19.3380 | 3.483 |
| State_NY | 19.5042 | 3.457 |
| State_OH | 19.5297 | 3.480 |
| State_OK | 19.1450 | 3.492 |
| State_OR | 18.4967 | 3.472 |
| State_PA | 19.6612 | 3.476 |
| State_RI | 19.3154 | 3.486 |
| State_SC | 19.6827 | 3.473 |
| State_SD | 18.5425 | 3.487 |
| State_TN | 19.2969 | 3.475 |
| State_TX | 20.1672 | 3.453 |
| State_UT | 18.9864 | 3.480 |
| State_VA | 19.5268 | 3.485 |
| State_VT | 17.4780 | 3.481 |
| State_WA | 19.0949 | 3.470 |
| State_WI | 19.1108 | 3.479 |
| State_WV | 18.4304 | 3.494 |
| State_WY | 17.2778 | 3.492 |
| t0 | -7.4256 | 1.630 |
| t2 | 0.8685 | 0.191 |
| b1 | 0.8611 | 0.026 |
| a1 | 0.6316 | 0.119 |
| a2 | 0.4650 | 0.075 |
| R-square | 0.9669 |  |
| Root MSE | 9.2939 |  |
| Observation | 4567 |  |

**Model 14**
| Variable | Coefficien | S.E. |
| --- | --- | --- |
| State_AK | 9.5460 | 9.9804 |
| State_AL | 11.9991 | 9.6295 |
| State_AR | 11.1770 | 9.7777 |
| State_AZ | 12.4550 | 9.5622 |
| State_CA | 12.2251 | 9.6924 |
| State_CO | 12.1578 | 9.4485 |
| State_CT | 12.6507 | 9.2511 |
| State_DC | 12.1266 | 9.3667 |
| State_DE | 12.0615 | 9.4223 |
| State_FL | 12.6138 | 9.5721 |
| State_GA | 12.3328 | 9.5209 |
| State_HI | 9.0987 | 10.0746 |
| State_IA | 11.8945 | 9.6341 |
| State_ID | 10.9464 | 9.7317 |
| State_IL | 12.4810 | 9.4842 |
| State_IN | 12.2584 | 9.4792 |
| State_KS | 11.3863 | 9.6608 |
| State_KY | 11.5100 | 9.6686 |
| State_LA | 12.5174 | 9.3058 |
| State_MA | 12.6781 | 9.3068 |
| State_MD | 12.3900 | 9.4628 |
| State_ME | 10.6601 | 9.6958 |
| State_MI | 12.4468 | 9.3113 |
| State_MN | 12.0540 | 9.6219 |
| State_MO | 11.6962 | 9.6647 |
| State_MS | 12.1683 | 9.5118 |
| State_MT | 10.0719 | 9.9861 |
| State_NC | 11.9679 | 9.7043 |
| State_ND | 10.8321 | 9.7155 |
| State_NE | 11.3572 | 9.7069 |
| State_NH | 11.6095 | 9.5981 |
| State_NJ | 12.9440 | 9.1431 |
| State_NM | 11.8357 | 9.6202 |
| State_NV | 11.9364 | 9.5677 |
| State_NY | 12.7671 | 9.1862 |
| State_OH | 12.1716 | 9.5718 |
| State_OK | 11.5981 | 9.6537 |
| State_OR | 10.9724 | 9.8202 |
| State_PA | 12.5207 | 9.4291 |
| State_RI | 12.2865 | 9.3524 |
| State_SC | 12.1198 | 9.6376 |
| State_SD | 10.9780 | 9.7599 |
| State_TN | 11.6459 | 9.7493 |
| State_TX | 12.5037 | 9.7180 |
| State_UT | 11.2319 | 9.8406 |
| State_VA | 12.1622 | 9.5705 |
| State_VT | 10.0472 | 9.5578 |
| State_WA | 11.9331 | 9.5323 |
| State_WI | 11.6491 | 9.6614 |
| State_WV | 10.7205 | 9.8812 |
| State_WY | 9.3713 | 9.9413 |
| t0 | -4.5443 | 3.9531 |
| t2 | 0.5808 | 0.4116 |
| c1 | -0.5533 | 0.3046 |
| c2 | 0.0326 | 0.0347 |
| b1 | 0.8778 | 0.0274 |
| w1 | 0.2925 | 0.0515 |
| w2 | 0.2981 | 0.0458 |
| R-square | 0.9679 |  |
| RMSE | 9.1536 |  |
| Observatio | 4567 |  |

**Model 15**
| Variable | Coefficien | S.E |
| --- | --- | --- |
| State_AK | 3.5952 | 8.7 |
| State_AL | 5.8859 | 8.4 |
| State_AR | 5.0183 | 8.5 |
| State_AZ | 6.3802 | 8.4 |
| State_CA | 6.0203 | 8.5 |
| State_CO | 6.1577 | 8.2 |
| State_CT | 6.8051 | 8.1 |
| State_DC | 6.2628 | 8.2 |
| State_DE | 6.1560 | 8.2 |
| State_FL | 6.4954 | 8.4 |
| State_GA | 6.2742 | 8.3 |
| State_HI | 3.1812 | 8.8 |
| State_IA | 5.8288 | 8.4 |
| State_ID | 4.8809 | 8.5 |
| State_IL | 6.4442 | 8.3 |
| State_IN | 6.2457 | 8.3 |
| State_KS | 5.2980 | 8.4 |
| State_KY | 5.3820 | 8.4 |
| State_LA | 6.6533 | 8.1 |
| State_MA | 6.7704 | 8.1 |
| State_MD | 6.3927 | 8.3 |
| State_ME | 4.5793 | 8.5 |
| State_MI | 6.5401 | 8.1 |
| State_MN | 5.9745 | 8.4 |
| State_MO | 5.5684 | 8.4 |
| State_MS | 6.1664 | 8.3 |
| State_MT | 4.0863 | 8.7 |
| State_NC | 5.8276 | 8.5 |
| State_ND | 4.7759 | 8.5 |
| State_NE | 5.2583 | 8.5 |
| State_NH | 5.5735 | 8.4 |
| State_NJ | 7.0991 | 8.0 |
| State_NM | 5.8012 | 8.4 |
| State_NV | 5.8771 | 8.4 |
| State_NY | 6.8951 | 8.0 |
| State_OH | 6.0791 | 8.4 |
| State_OK | 5.4982 | 8.4 |
| State_OR | 4.8834 | 8.6 |
| State_PA | 6.5106 | 8.2 |
| State_RI | 6.4248 | 8.2 |
| State_SC | 6.0074 | 8.4 |
| State_SD | 4.8780 | 8.5 |
| State_TN | 5.4874 | 8.5 |
| State_TX | 6.2748 | 8.5 |
| State_UT | 5.1468 | 8.6 |
| State_VA | 6.0770 | 8.4 |
| State_VT | 3.9987 | 8.3 |
| State_WA | 5.8807 | 8.3 |
| State_WI | 5.5418 | 8.4 |
| State_WV | 4.6024 | 8.6 |
| State_WY | 3.3305 | 8.6 |
| t0 | -2.4685 | 3.3 |
| t2 | 0.3694 | 0.3 |
| c1 | -0.5198 | 0.2 |
| c2 | -0.0379 | 0.1 |
| c3 | -0.0058 | 0.0 |
| c4 | -0.0017 | 0.0 |
| b1 | 0.9300 | 0.0 |
| w1 | 0.2836 | 0.0 |
| w2 | 0.2764 | 0.0 |
| R-square | 0.9691 |  |
| RMSE | 8.9888 |  |
| Observations | 4567 |  |

**Model 16**
| Variable | Coefficien | S.E. |
| --- | --- | --- |
| State_AK | 17.0018 | 5.543 |
| State_AL | 18.7653 | 5.641 |
| State_AR | 17.7498 | 5.625 |
| State_AZ | 19.7100 | 5.603 |
| State_CA | 19.9473 | 5.569 |
| State_CO | 19.3527 | 5.685 |
| State_CT | 19.6768 | 5.632 |
| State_DC | 18.4803 | 5.665 |
| State_DE | 18.4964 | 5.664 |
| State_FL | 20.3097 | 5.558 |
| State_GA | 19.4903 | 5.624 |
| State_HI | 17.0232 | 5.663 |
| State_IA | 18.5713 | 5.661 |
| State_ID | 17.5102 | 5.660 |
| State_IL | 19.9904 | 5.622 |
| State_IN | 19.6328 | 5.666 |
| State_KS | 17.9009 | 5.678 |
| State_KY | 18.5673 | 5.670 |
| State_LA | 18.8248 | 5.563 |
| State_MA | 19.8956 | 5.587 |
| State_MD | 19.4440 | 5.640 |
| State_ME | 17.9484 | 5.674 |
| State_MI | 20.1710 | 5.625 |
| State_MN | 19.2714 | 5.640 |
| State_MO | 19.0063 | 5.619 |
| State_MS | 18.8587 | 5.626 |
| State_MT | 17.7958 | 5.606 |
| State_NC | 19.1364 | 5.612 |
| State_ND | 17.2322 | 5.626 |
| State_NE | 17.7791 | 5.673 |
| State_NH | 18.6044 | 5.691 |
| State_NJ | 20.1089 | 5.617 |
| State_NM | 18.6012 | 5.609 |
| State_NV | 18.5251 | 5.616 |
| State_NY | 20.0549 | 5.555 |
| State_OH | 19.7788 | 5.660 |
| State_OK | 17.6217 | 5.667 |
| State_OR | 17.9494 | 5.662 |
| State_PA | 20.2618 | 5.644 |
| State_RI | 18.5871 | 5.607 |
| State_SC | 18.9771 | 5.617 |
| State_SD | 17.2530 | 5.674 |
| State_TN | 18.2918 | 5.584 |
| State_TX | 20.2237 | 5.597 |
| State_UT | 17.5312 | 5.598 |
| State_VA | 19.2248 | 5.646 |
| State_VT | 17.3072 | 5.641 |
| State_WA | 18.8195 | 5.607 |
| State_WI | 18.6382 | 5.628 |
| State_WV | 17.7771 | 5.633 |
| State_WY | 15.9743 | 5.677 |
| t0 | -6.8242 | 2.693 |
| t2 | 0.5852 | 0.317 |
| n1 | -0.3077 | 0.084 |
| pc1 | 0.5128 | 0.243 |
| bp | 0.7638 | 0.055 |
| d3 | 0.2386 | 0.058 |
| R-square | 0.9406 |  |
| Root MSE | 12.4556 |  |
| Observations | 4567 |  |

**Model 17**
| Variable | Coefficien | S.E. |
| --- | --- | --- |
| State_AK | 44.3754 | 7.8 |
| State_AL | 45.0719 | 7.7 |
| State_AR | 45.0296 | 7.7 |
| State_AZ | 45.4897 | 7.7 |
| State_CA | 45.2471 | 7.8 |
| State_CO | 44.7237 | 7.7 |
| State_CT | 44.9710 | 7.8 |
| State_DC | 44.8480 | 7.7 |
| State_DE | 44.8834 | 7.7 |
| State_FL | 45.7236 | 7.8 |
| State_GA | 45.2116 | 7.7 |
| State_HI | 43.5360 | 7.5 |
| State_IA | 44.9209 | 7.7 |
| State_ID | 44.4862 | 7.6 |
| State_IL | 45.2043 | 7.8 |
| State_IN | 44.7918 | 7.7 |
| State_KS | 44.8676 | 7.6 |
| State_KY | 44.4355 | 7.7 |
| State_LA | 46.0003 | 7.9 |
| State_MA | 45.0429 | 7.8 |
| State_MD | 45.1639 | 7.8 |
| State_ME | 43.7347 | 7.6 |
| State_MI | 44.6835 | 7.7 |
| State_MN | 44.8802 | 7.7 |
| State_MO | 44.9420 | 7.7 |
| State_MS | 45.1696 | 7.7 |
| State_MT | 44.2548 | 7.6 |
| State_NC | 45.0362 | 7.7 |
| State_ND | 44.4879 | 7.7 |
| State_NE | 44.8758 | 7.7 |
| State_NH | 44.2717 | 7.7 |
| State_NJ | 45.2459 | 7.8 |
| State_NM | 45.0833 | 7.8 |
| State_NV | 45.1552 | 7.7 |
| State_NY | 45.2907 | 7.8 |
| State_OH | 44.6637 | 7.7 |
| State_OK | 44.7409 | 7.7 |
| State_OR | 43.9776 | 7.6 |
| State_PA | 44.8583 | 7.7 |
| State_RI | 44.8480 | 7.8 |
| State_SC | 45.0818 | 7.7 |
| State_SD | 44.4404 | 7.7 |
| State_TN | 45.3233 | 7.8 |
| State_TX | 45.3508 | 7.7 |
| State_UT | 44.9916 | 7.7 |
| State_VA | 45.0741 | 7.7 |
| State_VT | 43.1094 | 7.6 |
| State_WA | 44.5409 | 7.7 |
| State_WI | 44.7343 | 7.7 |
| State_WV | 44.1364 | 7.7 |
| State_WY | 43.7190 | 7.6 |
| nt0 | 0.3855 | 0.0 |
| mt | -0.0801 | 0.0 |
| t1 | -19.5476 | 3.7 |
| t2 | 2.3839 | 0.4 |
| nt7 | -0.1655 | 0.0 |
| b1 | 0.5856 | 0.0 |
| pc1 | -0.0096 | 0.0 |
| w1 | 0.5435 | 0.1 |
| w2 | 0.6873 | 0.0 |
| R-square | 0.9507 |  |
| Root MSE | 303.5030 |  |
| Observatio | 4592 |  |

**Model 18**
| Variable | Coefficien | S.E. |
| --- | --- | --- |
| L7.ReportedDailyD | 0.6986 | 0.006 |
| _cons | 5.0046 | 0.421 |
| R-square | 0.6893 |  |
| Root MSE | 26.5453 |  |
| Observations | 4639 |  |

**Model 19**
| Variable | Coefficien | S.E. |
| --- | --- | --- |
| L7.ReportedDailyD | 0.6681 | 0.006 |
| _cons | 6.6079 | 0.409 |
| R-square | 0.6820 |  |
| Root MSE | 25.8280 |  |
| Observations | 4639 |  |

**Model 20**
| Variable | Coefficien | S.E. |
| --- | --- | --- |
| L7.DailyDead | 0.7360 | 0.006 |
| _cons | 3.4596 | 0.386 |
| R-square | 0.7428 |  |
| Root MSE | 24.1541 |  |
| Observations | 4639 |  |

**Model 21**
| Variable | Coefficien | S.E. |
| --- | --- | --- |
| L7.DailyDead | 0.7854 | 0.003 |
| _cons | 3.1725 | 0.213 |
| R-square | 0.9148 |  |
| Root MSE | 13.3709 |  |
| Observations | 4639 |  |

**Model 22**
| Variable | Coefficient |
| --- | --- |
| State_AK | 0.2006 |
| State_AL | 1.5370 |
| State_AR | 0.8090 |
| State_AZ | 2.1963 |
| State_CA | 2.0034 |
| State_CO | 1.3761 |
| State_CT | 1.6062 |
| State_DC | 1.1951 |
| State_DE | 1.2206 |
| State_FL | 2.5509 |
| State_GA | 1.8229 |
| State_HI | 0.1409 |
| State_IA | 1.3681 |
| State_ID | 0.5918 |
| State_IL | 1.9454 |
| State_IN | 1.5884 |
| State_KS | 0.8553 |
| State_KY | 0.9743 |
| State_LA | 1.5894 |
| State_MA | 1.7707 |
| State_MD | 1.7541 |
| State_ME | 0.4285 |
| State_MI | 1.4539 |
| State_MN | 1.5664 |
| State_MO | 1.1551 |
| State_MS | 1.5599 |
| State_MT | 0.3378 |
| State_NC | 1.5993 |
| State_ND | 0.5339 |
| State_NE | 0.8839 |
| State_NH | 0.9883 |
| State_NJ | 1.7275 |
| State_NM | 1.2831 |
| State_NV | 1.3298 |
| State_NY | 1.5576 |
| State_OH | 1.6383 |
| State_OK | 1.0367 |
| State_OR | 0.6691 |
| State_PA | 1.8697 |
| State_RI | 1.3484 |
| State_SC | 1.7655 |
| State_SD | 0.6404 |
| State_TN | 1.2324 |
| State_TX | 2.7912 |
| State_UT | 0.9140 |
| State_VA | 1.6275 |
| State_VT | 0.2214 |
| State_WA | 1.2469 |
| State_WI | 1.0928 |
| State_WV | 0.5399 |
| State_WY | 0.1559 |

**Model 23**
| Variable | Coefficient | S.E. |
| --- | --- | --- |
| LogPopulatio | 0.3652 | 0.0430 |
| PercentBlack | 0.0197 | 0.0030 |
| PercentHispa | 0.0172 | 0.0049 |
| PercentAge8 | 0.1163 | 0.0627 |
| _cons | -2.6082 | 0.2842 |
| R-square | 0.7938 |  |
| Root MSE | 0.2845 |  |
| Observations | 51 |  |

**Model 24**
| Variable | Coefficient |
| --- | --- |
| State_AK | 30.8645 |
| State_AL | 236.4667 |
| State_AR | 124.4626 |
| State_AZ | 337.8853 |
| State_CA | 308.2178 |
| State_CO | 211.7076 |
| State_CT | 247.1032 |
| State_DC | 183.8660 |
| State_DE | 187.7866 |
| State_FL | 392.4490 |
| State_GA | 280.4530 |
| State_HI | 21.6738 |
| State_IA | 210.4832 |
| State_ID | 91.0498 |
| State_IL | 299.2881 |
| State_IN | 244.3758 |
| State_KS | 131.5789 |
| State_KY | 149.8908 |
| State_LA | 244.5222 |
| State_MA | 272.4093 |
| State_MD | 269.8608 |
| State_ME | 65.9177 |
| State_MI | 223.6777 |
| State_MN | 240.9784 |
| State_MO | 177.7016 |
| State_MS | 239.9922 |
| State_MT | 51.9721 |
| State_NC | 246.0431 |
| State_ND | 82.1384 |
| State_NE | 135.9798 |
| State_NH | 152.0495 |
| State_NJ | 265.7632 |
| State_NM | 197.3949 |
| State_NV | 204.5898 |
| State_NY | 239.6327 |
| State_OH | 252.0446 |
| State_OK | 159.4940 |
| State_OR | 102.9358 |
| State_PA | 287.6393 |
| State_RI | 207.4532 |
| State_SC | 271.6207 |
| State_SD | 98.5245 |
| State_TN | 189.5980 |
| State_TX | 429.4077 |
| State_UT | 140.6124 |
| State_VA | 250.3867 |
| State_VT | 34.0594 |
| State_WA | 191.8292 |
| State_WI | 168.1248 |
| State_WV | 83.0635 |
| State_WY | 23.9773 |

**Model 25**
| Variable | Coefficient |
| --- | --- |
| State_AK | 0.1660 |
| State_AL | 1.2716 |
| State_AR | 0.6693 |
| State_AZ | 1.8169 |
| State_CA | 1.6574 |
| State_CO | 1.1384 |
| State_CT | 1.3288 |
| State_DC | 0.9887 |
| State_DE | 1.0098 |
| State_FL | 2.1103 |
| State_GA | 1.5081 |
| State_HI | 0.1165 |
| State_IA | 1.1318 |
| State_ID | 0.4896 |
| State_IL | 1.6094 |
| State_IN | 1.3141 |
| State_KS | 0.7075 |
| State_KY | 0.8060 |
| State_LA | 1.3149 |
| State_MA | 1.4648 |
| State_MD | 1.4511 |
| State_ME | 0.3545 |
| State_MI | 1.2028 |
| State_MN | 1.2958 |
| State_MO | 0.9556 |
| State_MS | 1.2905 |
| State_MT | 0.2795 |
| State_NC | 1.3231 |
| State_ND | 0.4417 |
| State_NE | 0.7312 |
| State_NH | 0.8176 |
| State_NJ | 1.4291 |
| State_NM | 1.0615 |
| State_NV | 1.1001 |
| State_NY | 1.2886 |
| State_OH | 1.3553 |
| State_OK | 0.8577 |
| State_OR | 0.5535 |
| State_PA | 1.5467 |
| State_RI | 1.1155 |
| State_SC | 1.4606 |
| State_SD | 0.5298 |
| State_TN | 1.0195 |
| State_TX | 2.3091 |
| State_UT | 0.7561 |
| State_VA | 1.3464 |
| State_VT | 0.1831 |
| State_WA | 1.0315 |
| State_WI | 0.9041 |
| State_WV | 0.4467 |
| State_WY | 0.1289 |

**Model 26**
| Variable | Coefficient | S.E. |
| --- | --- | --- |
| State_AK | 19.1718 | 5.5455 |
| State_AL | 22.8740 | 5.5486 |
| State_AR | 22.0474 | 5.5568 |
| State_AZ | 23.5269 | 5.5377 |
| State_CA | 23.6248 | 5.5105 |
| State_CO | 22.8014 | 5.5331 |
| State_CT | 23.1423 | 5.5123 |
| State_DC | 22.3685 | 5.5664 |
| State_DE | 22.3263 | 5.5622 |
| State_FL | 23.5596 | 5.5434 |
| State_GA | 23.2117 | 5.5427 |
| State_HI | 19.7769 | 5.6115 |
| State_IA | 22.5782 | 5.5493 |
| State_ID | 20.8793 | 5.5795 |
| State_IL | 23.5277 | 5.5071 |
| State_IN | 23.0678 | 5.5314 |
| State_KS | 21.8978 | 5.5721 |
| State_KY | 22.2463 | 5.5515 |
| State_LA | 23.1644 | 5.5624 |
| State_MA | 23.3854 | 5.4903 |
| State_MD | 23.1839 | 5.5355 |
| State_ME | 21.1217 | 5.5520 |
| State_MI | 23.1641 | 5.5079 |
| State_MN | 22.9096 | 5.5305 |
| State_MO | 22.6315 | 5.5517 |
| State_MS | 22.8856 | 5.5500 |
| State_MT | 20.5095 | 5.5764 |
| State_NC | 23.0000 | 5.5447 |
| State_ND | 21.0049 | 5.5624 |
| State_NE | 21.9977 | 5.5622 |
| State_NH | 22.1678 | 5.5486 |
| State_NJ | 23.7615 | 5.5012 |
| State_NM | 22.4717 | 5.5759 |
| State_NV | 22.4211 | 5.5878 |
| State_NY | 23.7318 | 5.4834 |
| State_OH | 23.1541 | 5.5151 |
| State_OK | 22.1275 | 5.5729 |
| State_OR | 21.6158 | 5.5637 |
| State_PA | 23.4885 | 5.5143 |
| State_RI | 22.5008 | 5.5332 |
| State_SC | 22.7793 | 5.5456 |
| State_SD | 21.4924 | 5.5670 |
| State_TN | 22.5039 | 5.5482 |
| State_TX | 23.4079 | 5.5367 |
| State_UT | 21.9073 | 5.5616 |
| State_VA | 22.9497 | 5.5357 |
| State_VT | 20.5659 | 5.5622 |
| State_WA | 22.4466 | 5.5406 |
| State_WI | 22.2606 | 5.5282 |
| State_WV | 21.2802 | 5.5830 |
| State_WY | 19.8045 | 5.5516 |
| t0 | -7.7761 | 2.6721 |
| t2 | 0.8279 | 0.3255 |
| b1 | 0.5529 | 0.0634 |
| w1 | 0.3298 | 0.0830 |
| w2 | 0.5021 | 0.0666 |
| R-square | 0.8082 |  |
| Root MSE | 22.9496 |  |
| Observations | 4565 |  |

## Notes

### Competing Interest Statement

The authors have declared no competing interest.

### Clinical Trial

Epidemiology study based on publicly available (non-individually identified) data.

### Funding Statement

The authors declare no competing interests. The authors alone, and not their respective institutions, are responsible for the content of this paper. The contributions to this paper by S.L. Carson and T.K. Dye were done outside of their usual employment.

### Author Declarations

No IRB approval required as only publicly available (non-individual identifiable) data (e.g., daily state-level COVID deaths) used.

